# The benefits, harms, and cost-effectiveness of age-based and risk-stratified screening for prostate cancer with MRI

**DOI:** 10.64898/2025.11.28.25341221

**Authors:** Thomas Callender, Caroline M Moore, Elena Frangou, Mark Emberton, Nora Pashayan

**Affiliations:** Department of Public Health and Primary Care, University of Cambridge, Cambridge, UK; Division of Surgical and Interventional Science, University College London; Department of Urology, University College London Hospitals Trust; UCL Innovative Clinical Trials Unit, University College London

**Author notes:** Correspondence to: Dr Thomas Callender and Professor Nora Pashayan, Department of Public Health and Primary Care, Strangeways Research Laboratory, University of Cambridge Cambridge, CB1 8RN.

## Abstract

**Background:** Prostate cancer screening remains contested. MRI-first screening may reduce overdiagnosis by detecting fewer insignificant cancers than PSA-based strategies. However, the benefits, harms, and cost-effectiveness of MRI-first prostate cancer screening remain uncertain.

**Methods:** We simulated age-based and risk-stratified MRI-first screening strategies. Risk-stratification involved either a polygenic risk score or a PSA test, with a PSA threshold of 1ng/ml or 3ng/ml considered positive. We conducted extensive scenario and sensitivity analyses to determine the robustness of conclusions to input parameters. Primary outcomes were prostate cancer deaths, overdiagnosed cancers, resource use, and cost-effectiveness (net health benefit from a healthcare perspective).

**Results:** Age-based MRI-first screening every 4 years from 55–69 years old could reduce prostate cancer deaths by 6 per 1,000 men, with 13% of screen-detected cancers overdiagnosed. However, this strategy required a 15-fold increase in MRI scans relative to no screening and was not cost-effective within a UK context. Risk-stratified screening using polygenic risk was more likely to be cost-effective, generate more QALYs, and lead to fewer overdiagnosed cancers than screening all men with MRI. At a willingness-to-pay (WTP) of £25,000 per QALY gained, screening men aged 55 with a 10-year absolute risk of prostate cancer of ≥5.6% had a greater NHB than no screening. Stratifying screening with PSA before MRI led to 6%–17% more overdiagnosed cancers than MRI-first screening, depending on the threshold for a positive PSA test (3ng/ml or 1ng/ml, respectively). But PSA stratification was not cost-effective at either a 1ng/ml or 3ng/ml threshold in a UK context.

**Conclusions:** MRI-first screening could reduce overdiagnosis and generate more QALYs than stratifying screening with PSA prior to MRI. But the additional resource requirements meant that risk-stratification is likely to be necessary for prostate cancer screening to be considered cost-effective in the UK.

## Introduction

Prostate cancer is a leading cause of cancer death in men^1^. Screening for prostate cancer every four years with prostate-specific antigen (PSA) has been shown to reduce prostate-cancer-specific mortality by 13% (95% confidence interval [5%–20%]) after 23-years follow-up^2^. But the potential harms, particularly overdiagnosis, have hampered the development of a population-based screening programme. These harms include false positives, with only one-quarter of biopsies showing cancer^2^ and, in particular, overdiagnosis. Overdiagnosis – the detection of cancers through screening that would not otherwise have been diagnosed^3^ – drives overtreatment and subsequent treatment-related harms, reducing the overall benefits of screening^4^.

Screening for prostate cancer using magnetic resonance imaging (MRI) may have the potential to mitigate overdiagnosis without a loss of clinical effectiveness. When used as a triage test before biopsy following an elevated PSA, MRI (multiparametric or biparametric MRI) reduces the detection of clinically insignificant cancers, defined as Gleason score 6, in both diagnostic^5,6^ and screening^7–12^ settings without compromising the detection of clinically significant cancer. Preliminary studies using MRI as a first-line test for population screening – the Imperial Prostate 1 Prostate Cancer Screening Trial (IP1-PROSTAGRAM)^13^ and ReIMAGINE^14^ – suggest that these advantages may also hold in a screening setting. Enriching the screened cohort through prior risk-stratification may further improve the benefit-to-harm profile and cost-effectiveness of prostate cancer screening whilst reducing the resources needed^15^.

A prospective, multi-arm, randomised trial of MRI-first screening including polygenic risk-stratified arm has been launched^16^. Modelling can be used to analyse multiple alternative prostate cancer screening approaches under varying assumptions and thus inform strategies for further prospective evaluation. In this work, we aimed to estimate the potential impact and cost-effectiveness of MRI-first screening, with or without risk-stratification using polygenic risk or PSA, relative to no screening.

## Methods

### Model structure

We simulated cohorts of 1,000 men between the ages of 55 and 89 years. Using a life-table approach, we estimated the age-specific incidence of prostate cancer, deaths from prostate cancer, and deaths from other causes^15,17^. On this background, we superimposed alternative screening approaches both with and without prior risk stratification (see Appendix A for more details).

### Modelled scenarios

In our primary analyses, we compared no screening with MRI-first screening of men from the age of 55 to 69 at four-yearly intervals with MRI. In secondary analyses, we considered risk-stratified screening using either polygenic risk or a PSA test. The scenarios can be summarised as follows:

- *MRI-first screening*: Biparametric MRI (bpMRI) used as a first-line screening test, with a positive screen being a score of ≥ 4 using the Prostate Imaging-Reporting and Data System (PI-RADS) version 2. If positive, the man would be offered a diagnostic biopsy.
- *Risk-stratified screening*:

`o` **Polygenic risk:** Men first offered a polygenic risk score to determine – in combination with their age – their eligibility for screening. MRI screening, as described above, would then be offered to those over a risk threshold.
`o` **PSA:** PSA used as the first-line screening triage test, with thresholds of 1ng/ml or 3ng/ml considered a screen positive. Men over these thresholds would have a reflex bpMRI, with a PI-RADS score of ≥ 4 triggering referral for a diagnostic biopsy.

### Model parameters

Age-specific prostate cancer incidence, mortality from prostate cancer and mortality from other causes amongst men in England in the absence of screening were calculated from mean 2017-2019 Office for National Statistics and NHS England data^18,19^.

The clinical impact of screening depends on the combination of mean sojourn time (MST), episode sensitivity, and relative mortality reduction expected. The MST is the average time that a cancer is screen-detectable before clinical presentation^20^. Episode sensitivity describes the sensitivity of the entire diagnostic process, from a positive screening test result through to prostate cancer diagnosis^21^. The combination of MST and episode sensitivity determine the number of cancer diagnoses brought forwards by screening, interval cancers, and cancers overdiagnosed^22^. In our primary analyses of MRI-first screening:

- The relative mortality reduction with screening was 0.82 (95% CI: 0.77–0.86). Based on the findings of the ERSPC^2^ trial adjusted for the proportion of clinically significant cancers detected relative to PSA screening with a threshold of 3ng/ml.
- The MST was 7.6 (95% CI: 6.5–8.9) years.
- The sensitivity of a screening episode was set to 80% (95% CI: 70%–90%).

Model inputs and assumptions are presented in Appendix A (Tables A.1-A.5). To validate the reliability of our model’s performance, we compared it against the European Randomized Study of Screening for Prostate Cancer (ERSPC) (Figures B.1 in Appendix B).

### Scenario and sensitivity analyses

Given the uncertainty in key parameters and to allow for the wide number of potential screening pathways that might be used, we ran scenario analyses, varying each of the following variables: relative mortality reduction with screening; MST; episode sensitivity, proportion of screening tests that were positive and, the costs of screening MRI scans, biopsies, and polygenic risk stratification.

In each scenario, we re-ran the analyses, holding the parameter of interest constant at a specified value whilst continuing to draw all other parameters from their relevant distributions. Subsequently, we simultaneously varied the MST, the relative mortality reduction with screening, the proportion of screening tests positive, and the costs of screening bpMRI scans and biopsies to analyse the impact of different combinations of these five variables. We then analysed screening from ages 50-69, 50-74, and 55-74. Finally, we performed one-way sensitivity analyses of resource use and utility estimates^23^.

### Polygenic risk

We modelled the impact of stratified screening by centile of the polygenic risk distribution of prostate cancer. The polygenic risk score follows a log-normal relative risk distribution^24^, whose variance, given known prostate cancer susceptibility loci, is 0.72^25^. Using this distribution, we determined the proportion of prostate cancers, and hence the relative risk of cancer diagnosis amongst those above and below each absolute risk threshold. The average 10-year absolute risk of prostate cancer at age 55 is 2.6% (Figure A.1 in Appendix A). By multiplying this value by the centile-specific relative risk of cancer (Table A.6 in Appendix A), we determined the 10-year absolute risk by centile of the polygenic distribution.

### Model outputs

We calculated outcomes including prostate cancer diagnoses, deaths from prostate cancer and other causes, PSA tests, biopsies, screening and diagnostic MRI, as well as life-years, quality-adjusted life-years (QALYs) and costs. We calculated the proportion of screen-detected cancers overdiagnosed using the lead time approach (see Appendix A methods for more details)^20^.

### Cost-effectiveness analyses

We calculated the net health benefit (NHB) of different scenarios. NHB considers health gains (QALYs) in the context of the opportunity cost of those foregone in the population by introducing a particular intervention, derived as *NHB_i_ = QALY_i_ – (Cost_i_ / threshold)*, where *i* is the intervention or strategy and the willingness-to-pay (WTP) threshold for one QALY^26^. Net health benefit is directly proportional to net monetary benefit. We considered UK National Institute for Health and Care Excellence (NICE) thresholds of £25,000 and £35,000 per QALY gained in our main analyses^27^. The strategy with the highest NHB at a particular willingness-to-pay threshold was considered optimal. Future costs and benefits were both discounted by 3.5%^28^. Utility estimates, resource use, and cost estimates are presented in Appendix A (Tables A.2 and A.7-A.10).

### Statistical analyses

All reported analyses are probabilistic: we ran 10,000 simulations for each scenario studied, drawing parameters simultaneously from their distributions. For intervals, we present the 2.5^th^ and 97.5^th^ centiles of the sorted probabilistic results.

## Code and data availability

The data and code used in these analyses are openly available at https://github.com/callta/mri-screening-prostate. We used Devcan version 6.7.8.5^29^ and Julia^30^ version 1.11.6 for analyses. We adhered to the Consolidated Health Economic Evaluation Reporting Standards (CHEERS) reporting guideline^31^; a completed checklist is available as an Appendix.

## Results

### Age-based MRI-first screening

Compared with no screening, age-based MRI-first screening reduced deaths from prostate cancer by 6.1 (95% interval: 4.8–7.5) per 1,000 men; a relative reduction of 18.2% (95% interval: 14.3%–22.4%) (Table 1, Appendix B Table B.1). The absolute risk difference was 0.30% (95% interval: 0.23%–0.37%) after 25 years follow-up of the cohort from age 55.

**Table 1:** Outcomes per 1,000 men screened with age-based or risk-stratified MRI screening from age 55 every 4 years to 69.

| Strategy | Prostate cancers | Screen-detected cancers | Interval cancers | Overdiag-nosed cancers | Deaths from prostate cancer | Relative reduction in prostate cancer deaths (%) | bpMRI | mpMRI | Biopsies | Life-years | QALYs | Costs (GBP £) | NHB (QALYs) |
| --- | --- | --- | --- | --- | --- | --- | --- | --- | --- | --- | --- | --- | --- |
| No screening | 140 | 0 | 0 | 0 | 34 | 0 | 0 | 263 | 176 | 16,965 | 14,211 | 878,516 | 14,175 |
| MRI-first screening | 149 | 57 | 13 | 8 | 27 | 18 | 3,693 | 172 | 507 | 16,984 | 14,219 | 1,550,738 | 14,157 |
| *Polygenic risk-stratified screening* |  |  |  |  |  |  |  |  |  |  |  |  |  |
| 10th centile | 148 | 56 | 12 | 7 | 27 | 18 | 3,313 | 172 | 467 | 16,984 | 14,219 | 1,534,128 | 14,158 |
| 20th centile | 146 | 54 | 12 | 7 | 27 | 18 | 2,934 | 173 | 427 | 16,984 | 14,219 | 1,458,808 | 14,161 |
| 30th centile | 145 | 51 | 11 | 7 | 28 | 18 | 2,557 | 176 | 389 | 16,983 | 14,219 | 1,384,260 | 14,164 |
| 40th centile | 144 | 48 | 11 | 6 | 28 | 17 | 2,182 | 180 | 352 | 16,983 | 14,219 | 1,310,478 | 14,167 |
| 50th centile | 143 | 44 | 10 | 6 | 28 | 17 | 1,808 | 184 | 315 | 16,982 | 14,219 | 1,237,582 | 14,170 |
| 60th centile | 141 | 39 | 9 | 5 | 28 | 16 | 1,437 | 191 | 280 | 16,980 | 14,219 | 1,165,836 | 14,172 |
| 70th centile | 140 | 34 | 7 | 4 | 29 | 14 | 1,068 | 199 | 247 | 16,979 | 14,218 | 1,095,774 | 14,174 |
| 80th centile | 138 | 26 | 6 | 3 | 30 | 12 | 702 | 211 | 216 | 16,976 | 14,217 | 1,028,573 | 14,176 |
| 90th centile | 137 | 16 | 4 | 2 | 31 | 8 | 343 | 227 | 189 | 16,973 | 14,216 | 967,618 | 14,177 |
| PSA (1ng/ml) | 150 | 58 | 14 | 9 | 28 | 17 | 1,290 | 173 | 392 | 16,982 | 14,216 | 1,269,439 | 14,165 |
| PSA (3ng/ml) | 150 | 52 | 16 | 8 | 29 | 15 | 621 | 184 | 235 | 16,980 | 14,215 | 1,074,011 | 14,172 |

Net health benefits were calculated at a willingness-to-pay threshold of £25,000 per QALY gained. Net health benefits are calculated as $qalys - (costs / threshold)$ . Net health benefits are proportional to net monetary benefit $[(threshold * QALYs) - costs]$ . Strategies with a greater NHB than no screening at a given threshold produce more net QALYs. 1ng/ml and 3ng/ml refer to thresholds for considering a PSA test as positive. Life-years, QALYs, costs, and resulting NHB use discounted values (discount rate of 3.5%). Reduction in prostate cancer deaths is relative to no screening.
\* Numbers appear equal due to rounding.
Abbreviations: MRI, magnetic resonance imaging; bpMRI, biparametric MRI; mpMRI, multiparametric MRI; QALYs, quality-adjusted life-years; NHB, net health benefit; PSA, prostate specific antigen.

There were 7.6 overdiagnosed cancers per 1,000 men (95% interval: 5.5–10.2), representing 13.3% (95% interval: 10.8%–16.0%) of the 57 (95% interval: 48–67) screen-detected cancers (Appendix B Table B.2). This equated to 1.3 (95% interval: 0.9–1.8) overdiagnosed cancers per cancer death prevented. The overall rise in prostate cancer incidence with age-based MRI-first screening was 5.9% (95% interval: 0.8% –11.7%).

MRI-first screening led to a 15-fold rise in imaging requirements relative to no screening. The total number of MRI scans rose from 263 (95% interval: 176–353) to 3,865 (95% interval: 3,801–3,931) per 1,000 men, whilst there was a 3-fold increase in the number of biopsies from 176 (95% interval: 114–247) to 507 (95% interval 395–631).

By comparison with no screening, MRI-first screening led to a gain in 7.9 QALYs (95% interval: –0.3 to 14.9) for an additional £672,222 (95% interval: £299,982–£1.2M) per 1,000 men (Figure 1). The probability that age-based MRI screening had a greater NHB than no screening at willingness-to-pay (WTP) thresholds of £25,000 and £35,000 per QALY gained was 1.7% and 6.7%, respectively (Figure 2). A WTP threshold of £82,000 was necessary for age-based MRI-first screening to be more cost-effective than no screening.

**Figure 1:**
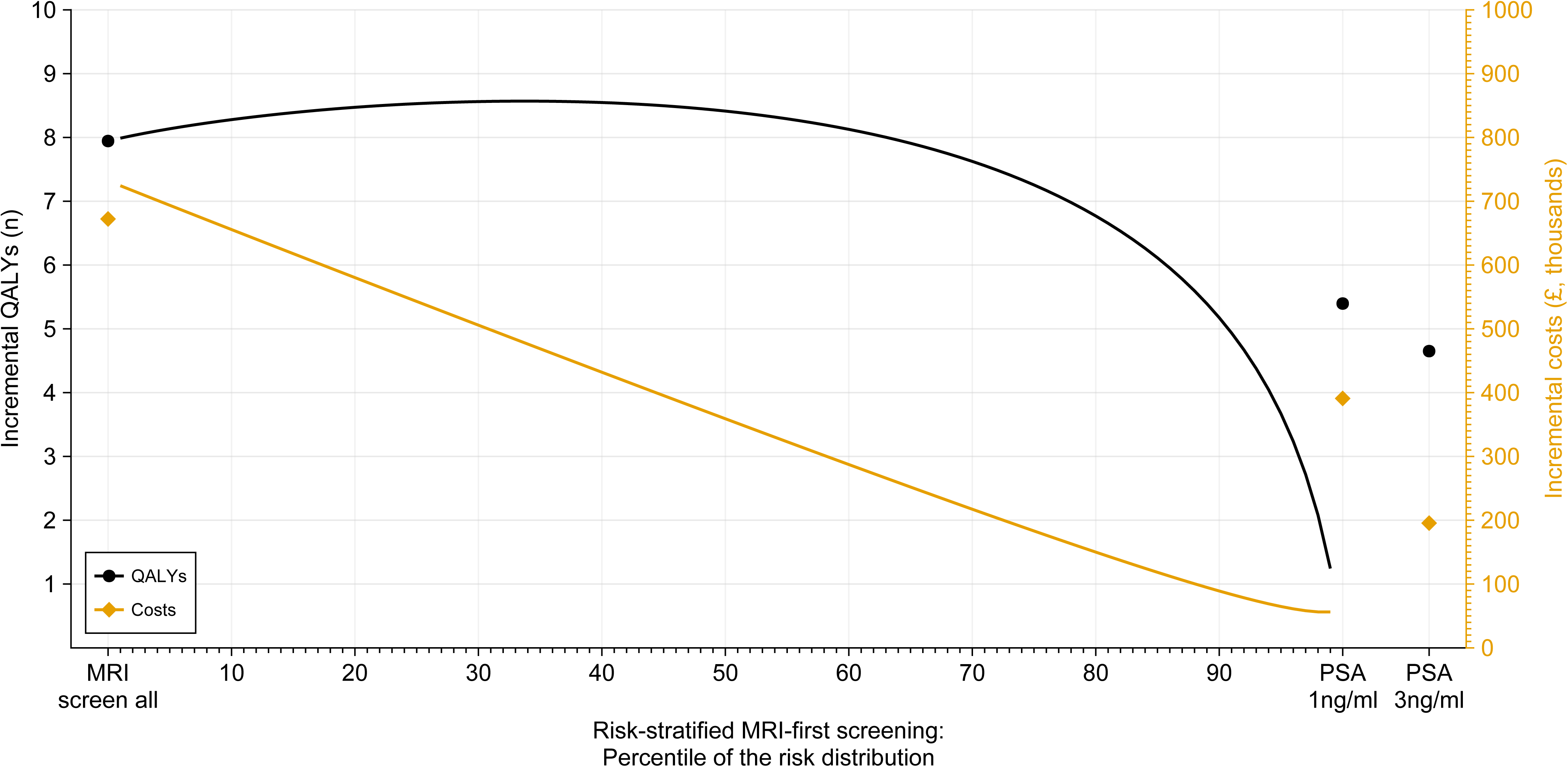
Expected costs (blue lines) and QALYs (black lines) relative to no screening per 1,000 men screened from age 55-69 every four years with age-based (“Screen all”) and polygenic risk-stratified MRI-first strategies. The highest number of incremental QALYs were generated when screening men above the 30^th^–37^th^ centiles.

**Figure 2:**
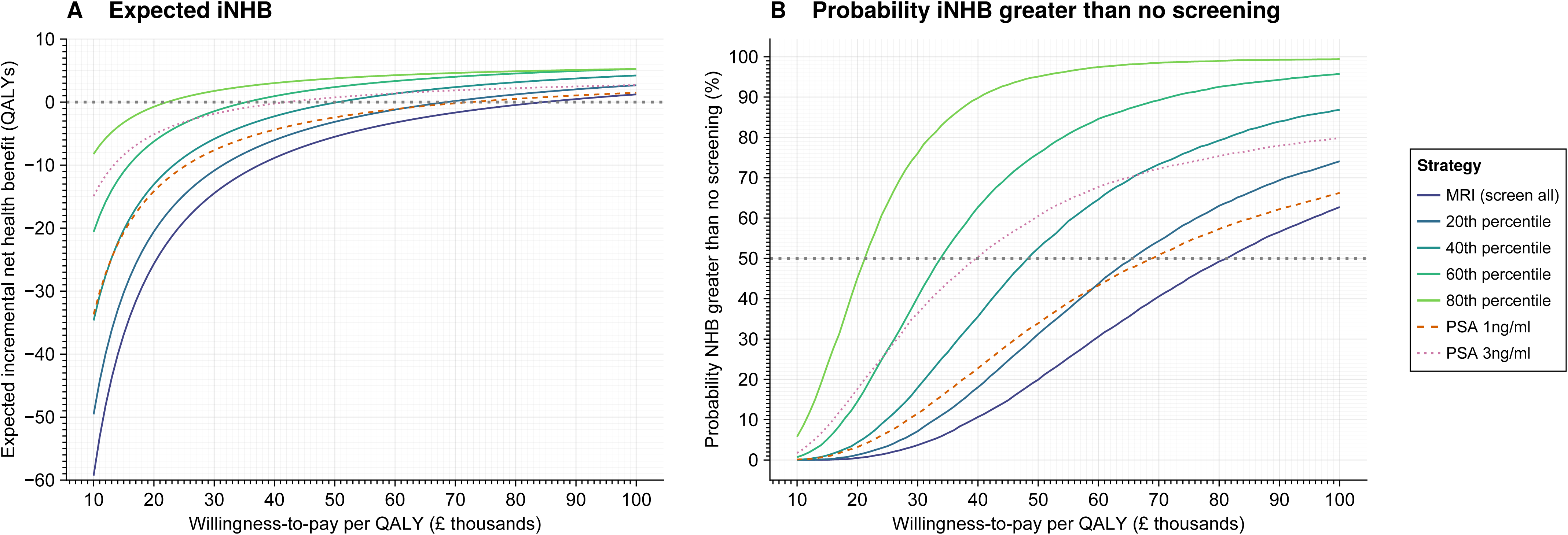
Net health benefit (NHB) by willingness-to-pay (WTP) threshold. (A) shows the expected incremental NHB of MRI-first screening relative to no screening, alongside (B) the probability of the incremental NHB being greater than that of no screening. The probability of having an incremental NHB > 0 provides a measure of the spread of outcomes relative to no screening. For example, at a WTP of £25,000 per quality-adjusted life-year (QALY), age-based MRI-first screening had an incremental NHB of -19 QALYs (subfigure A), with a 1.7% probability of being more cost-effective than no screening (subfigure B). At each WTP threshold, the strategy with the highest NHB is the most cost-effective.

### The impact of risk-stratification using polygenic risk

Risk-stratified strategies between the 7^th^ and the 64^th^ centile of the polygenic risk distribution generated more QALYs and had fewer costs than screening all men (Figure 1 & Appendix B Table B.1 and B.3). The greatest number of QALYs (14,219) were accrued by screening men from the 34^th^ centile (≥3.5% 10-year absolute risk of prostate cancer). More selective risk-stratified screening strategies had fewer resource requirements and were more cost-effective than screening all men, but at the expense of fewer prostate cancer deaths prevented (Table 1; Appendix B Tables B.1 and Appendix A Table A.6). Between the 10^th^ and the 90^th^ centiles, the number overdiagnosed reduced from 7.5 to 2.1 per 1,000 – reflecting 13% of screen-detected cases across these strategies – whilst the relative number of deaths prevented reduced from 18% to 8%.

At a WTP of £25,000 per QALY gained, screening men from the 74^th^ centile (≥5.6% 10-year absolute risk of prostate cancer) was more cost-effective than no screening (NHB: 14,175; probability incremental NHB [iNHB] greater than no screening: 50.5%). The MRI-first screening strategy with the highest NHB was more selective – screening only those from the 90^th^ centile of the polygenic risk distribution (NHB: 14,177; iNHB > no screening: 82.3%).

As the WTP threshold increased to £35,000, screening men from the 59^th^ centile (≥4.6% 10-year absolute risk of prostate cancer) had a higher NHB than no screening (NHB: 14,185; iNHB > no screening: 51.2%), with restricting screening to those above the 86^th^ centile being most cost-effective (NHB: 14,188; iNHB > no screening: 92.1%).

### The impact of stratifying screening based on a PSA test

PSA, used prior to MRI, led to fewer deaths prevented from prostate cancer and more overdiagnosis than MRI-first screening. PSA stratification with a threshold of 1ng/ml resulted in 5.8 (95% CI: 4.6–7.0) fewer deaths from prostate cancer per 1,000 men by comparison with no screening; a 17.1% reduction (95% CI: 13.6%–20.9%). 8.9 (95% CI: 6.2–12.1) men would be overdiagnosed per 1,000 – 15.2% of screen-detected cancers (95% CI: 12.2%– 18.4%). Raising the PSA threshold to 3ng/ml led to fewer cancer deaths prevented but lower overdiagnosis (Table 1).

PSA stratification generated fewer life-years and QALYs than MRI-first screening. However, PSA stratification-based strategies were less resource intensive, requiring fewer MRI and biopsies for expected mortality reductions. To have a higher NHB than no screening, stratified screening needed a WTP value of £69,000 when using a PSA threshold of 1ng/ml, and £40,000 at a PSA threshold of 3ng/ml (Figure 3).

**Figure 3:**
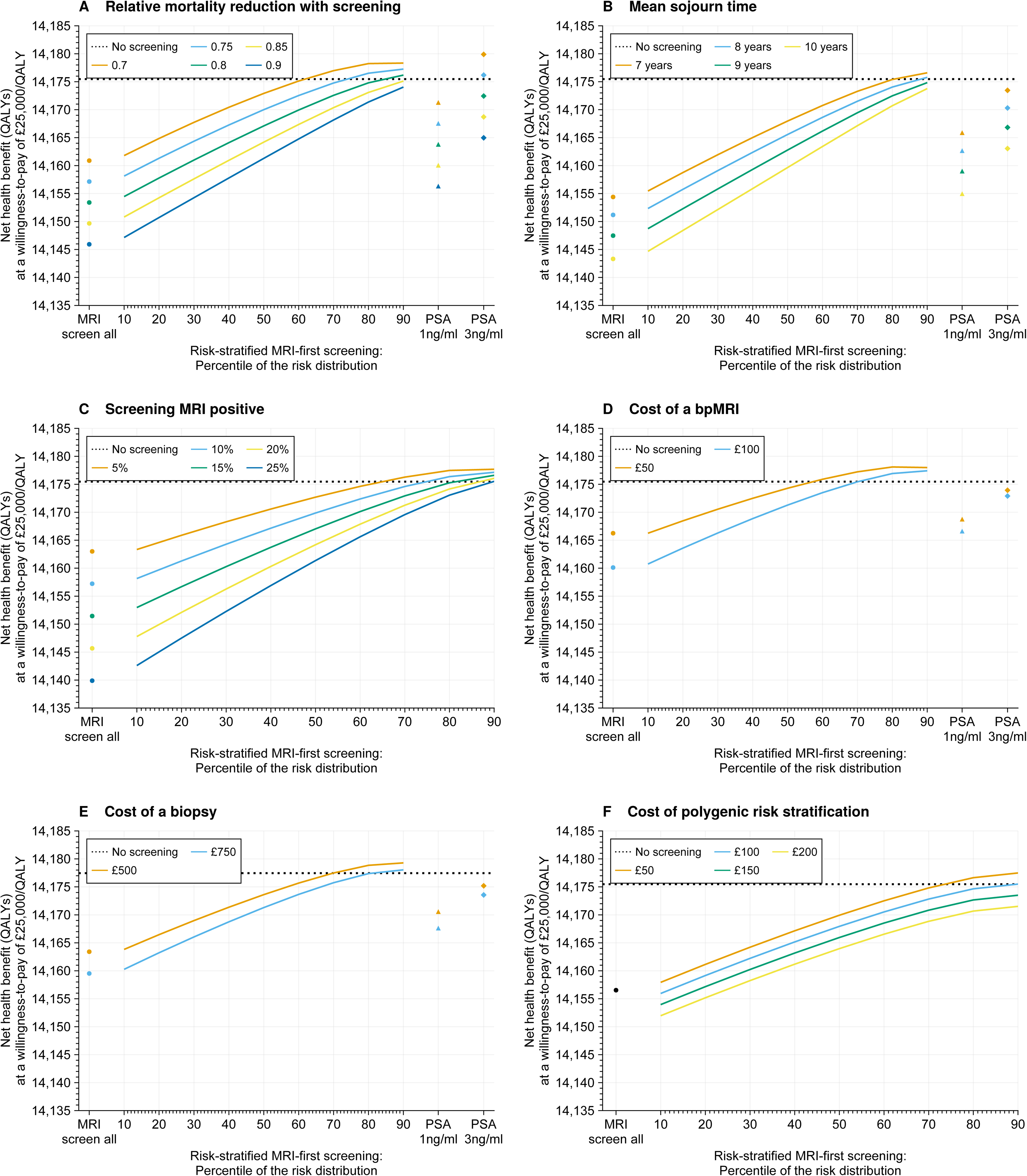
Scenario analyses: impact of alternative parameter values on the Net Health Benefit (NHB) of MRI-first screening at a willingness-to-pay (WTP) threshold of £25,000 per QALY gained. In all subplots, the dotted horizontal line reflects the NHB of no screening. If the NHB of screening is lower than that of no screening, this indicates the strategy – given the relevant parameter values – would not be cost-effective at this WTP threshold. Equivalent plots for MRI screening using a WTP threshold of £35,000 are presented in Appendix B.

### Scenario and sensitivity analyses

Greater relative mortality reductions and lower MSTs improved the cost-effectiveness of screening strategies (Figure 3 and Appendix B Figure B.3 & Tables B.4-B.5). Screening effectiveness is driven by earlier detection of a cancer – governed by the MST and episode sensitivity – coupled with the relative mortality reductions expected. With a shorter MST, screening anticipates fewer years of cancer diagnoses. In turn, this will reduce overdiagnosis and the number of life-years lived with prostate cancer. A shorter MST would be expected when a greater proportion of cancers detected by screening are clinically significant. As the proportion of cancers detected at a clinically significant (Gleason 7+) stage increases from 70% to 90%, the MST reduced from approximately 9.7 years to 7.5 years. Screening scenarios involving lower resource use – either through fewer MRI screening tests being positive for a given cancer detection rate or by a reduction in the costs of MRI – were more cost-effective, allowing for risk thresholds for eligibility to be further relaxed.

However, in none of our scenario analyses was age-based MRI-first screening cost-effective within a UK context. For an age-based MRI-first screening strategy to be cost-effective within a NICE threshold of £35,000 per QALY, the mortality reduction with screening must be <0.8, whilst the cost of a screening MRI would need to be reduced to less than £50 and that of a biopsy, £500, respectively (Figure 4 and Appendix B Figure B.3). Such values are approximately half of current cost estimates (Tables A.2 & A.10 in Appendix A).

**Figure 4:**
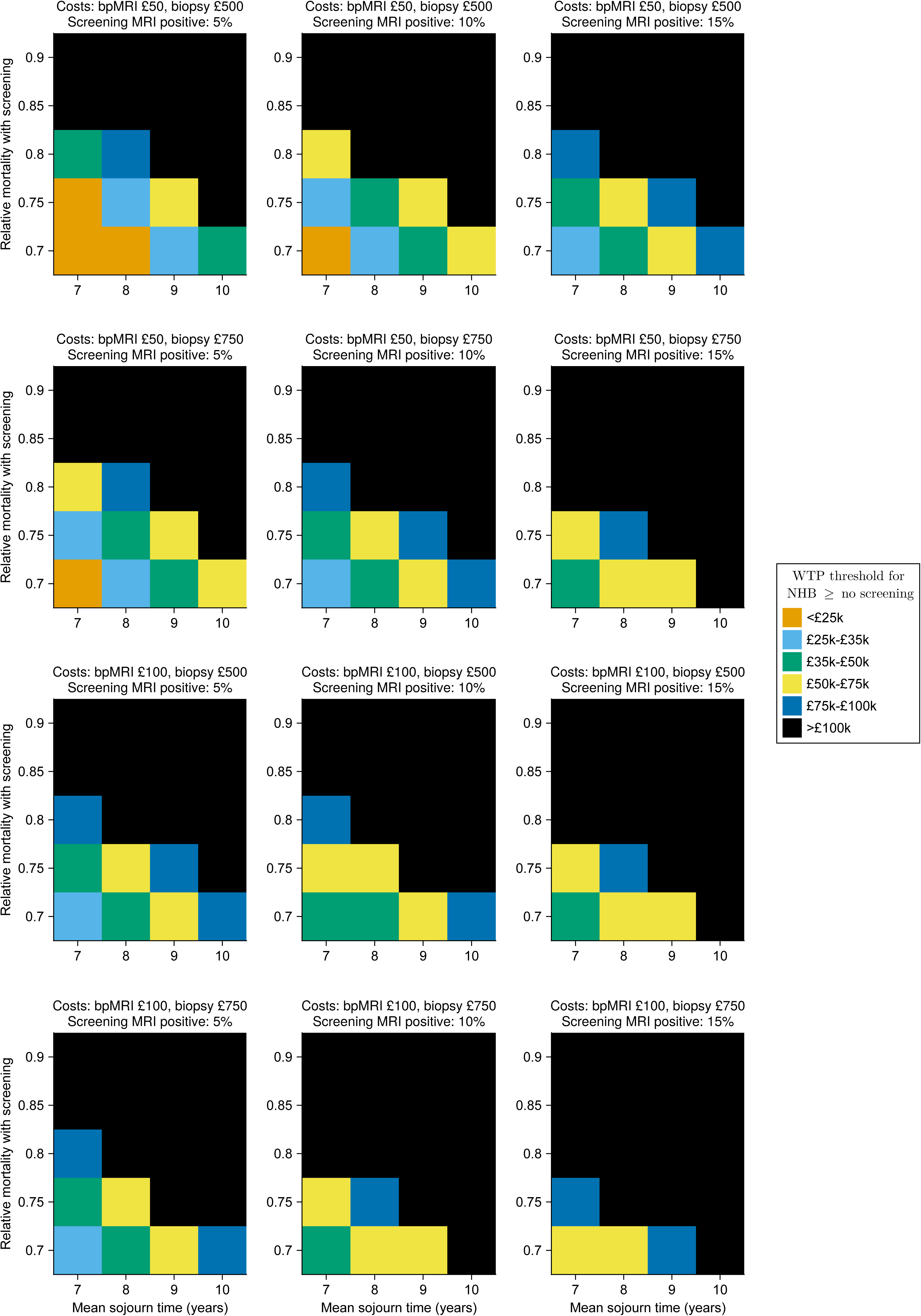
Threshold analyses: impact of simultaneously varying the relative mortality with screening, mean sojourn time (MST), proportion of screening tests that are positive, and the costs of biparametric screening MRIs (bpMRI) and biopsies. As an illustration, in the top left subpanel, with a relative mortality of 0.75 (y axis), an MST of 7 (x axis), 5% of screening MRIs positive, costs of bpMRI of £50 and that of biopsy £500, then MRI-first screening would have an incremental NHB greater than no screening at a WTP threshold of <£35,000 per QALY gained.

Lowering the age at which screening starts from 55 to 50, or increasing the age at which it stops from 69 to 74 reduced the cost-effectiveness of screening, whilst screening beyond 70 increased overdiagnosis and thus worsened the benefit/harm profile of screening (Appendix B Table B.6). MRI-first screening using a less restrictive threshold (PI-RADS ≥ 3) for screen positives led to greater numbers of diagnostic biopsies and proved less cost-effective (Appendix B Table B.7 and Figure B.4). Our model was insensitive to most costs except for those of biopsies and bpMRI (probabilistic one-way sensitivity analyses shown in Appendix B Figure B.5).

## Discussion

In this analysis, we show that risk-stratified prostate cancer screening strategies using a polygenic risk score or a PSA test could be both clinically- and cost-effective under a broad range of parameter assumptions.

Age-based screening with MRI as a primary screening test prevented the most prostate cancer deaths. However, the resources entailed meant this would not be cost-effective within a UK context. By contrast, stratifying the population prior to an MRI with a polygenic risk score accrued the most QALYs gained of the strategies compared and could be cost-effective in a UK context. We found that using PSA with either a threshold of 1ng/ml or 3ng/ml prior to a reflex bpMRI had a higher NHB than using MRI alone as a screening test within a UK context. This was because recent evidence from the Göteborg-2^32^ and STHLM-3^7^ studies has shown that the proportion of clinically significant screen-detected cancers with this screening strategy would be higher than previously estimated^15^. Nonetheless, stratifying screening using PSA generated fewer QALYs and had a lower NHB at both WTP thresholds of £25,000 and £35,000 per QALY gained than using a polygenic risk score.

The use of a polygenic risk score or PSA test to triage individuals have different implications for the cancers detected. We assume that a polygenic risk score will not differentiate between cancer types, leaving bpMRI to filter clinically insignificant cancers. Polygenic risk is thus used here as a predictive marker to determine absolute risk, with a threshold then applied to determine eligibility for screening. By contrast, PSA is a diagnostic marker which leads to a specific pool of potentially detectable cancers, acting as a prior filter on the cancers detectable before reflex bpMRI. Notably, 15% of prostate cancers occur in men with a PSA <3ng/ml, and amongst these cancers, 30% have a Gleason score of ≥ 7^33^. This highlights that a PSA-first strategy, in which only men with a PSA ≥ 3ng/ml undergo reflex bpMRI, would miss a substantial proportion of clinically significant disease. In IP1-PROSTAGRAM, the use of PSA with a threshold of 3ng/ml to trigger a reflex bpMRI was less sensitive for clinically significant cancer than either a 1ng/ml PSA threshold or screening all men with MRI (detection rates of clinically significant cancer of 1.5%, 2.5%, and 2.7%, respectively)^10^. Furthermore, 60% of those who screened positive with MRI and had a clinically significant cancer had a PSA <3ng/ml in the RE-IMAGINE study^14^. Correspondingly, as our analyses show, such a strategy could be expected to lead to more interval cancers and fewer prostate cancer deaths prevented than alternative screening approaches.

The resource intensity – MRI scans and, in particular, biopsies – of screening was a key driver of cost-effectiveness. We found that MRI strategies in which a positive screening MRI led directly to biopsy might lead to higher biopsy use than PSA screening. This was because biparametric MRI as a screening test led to a higher number screening positive in the IP1-PROSTAGRAM^13^ study than PSA screening did. True biopsy requirements remain uncertain because of the relatively small sample size of IP1-PROSTAGRAM. However, in threshold analyses, we show that MRI-first screening requires drops in both costs and resource intensity for such a programme to be likely to be cost-effective within plausible parameter estimates for the relative mortality benefit and MST of MRI-first screening (Figure 4).

We based our primary mortality analyses on the ERSPC as the only randomized trial to show evidence of prostate cancer mortality reduction with screening^2^. Extrapolating these results to MRI-first screening and PSA screening with a relaxed PSA threshold (1ng/ml vs 3ng/ml) required assumptions, of which estimates of sojourn time have proved most important as it underpin the advantages of MRI-first screening at a given relative mortality reduction. Our estimates of the MST were approximately 8 years for MRI-first, shorter than the approximately 9 years for PSA screening. This is due to a lower proportion of screen-detected cancers being clinically insignificant, with these Gleason grade 6 cancers estimated to have a more indolent natural history and longer sojourn time than clinically significant cancers. These assumptions are consistent with recent studies^7,8,11,34,35^ and would imply lower numbers of overdiagnosed cancers than in previous trials, such as ERPSC.

In keeping with the literature, we show that strategies using PSA followed by reflex bpMRI as a triage test could be cost-effective^36^. Furthermore, we also show that screening all men with an MRI-first approach could increase the number of biopsies required relative to a PSA followed by MRI approach and has a low probability of being cost-effective^37^. However, by contrast with Gulati and colleagues^37^, we did not find that overdiagnosis would be higher with age-based MRI-first than PSA screening, nor do we feel there are strong grounds to assume that MRI-first screening will necessarily lead to fewer deaths prevented than an equivalent PSA screening programme. Nevertheless, varying these values in our scenario analyses did not fundamentally change our shared conclusion.

This work has several strengths. We used a transparent simulation approach where key underpinning parameters could be directly inputted. This allowed us to explore a wide range of scenario and sensitivity analyses, particularly in key drivers of the clinical- and cost-effectiveness of any screening programme: relative mortality reduction, mean sojourn time, and episode sensitivity. We do not rely on assumptions of stage shift with regards relative mortality reduction which have been shown to be unreliable in prostate cancer screening^38^. We considered a range of scenarios and MRI pathways, including risk-stratified approaches to screening. We have made the underpinning analytical code open-source and have developed a web application to allow interactive analyses with different parameters and assumptions.

## Limitations

Prospective trials with long-term follow-up using MRI as the screening test are not available, such that the model is based on assumptions. We have tried to make these as explicit as possible. We have also undertaken extensive scenario analyses to explore how confident one can be in our conclusions. We constrained the number of screening strategies studied, choosing not to consider multiple different starting/stopping ages and screening intervals, or alternative approaches to risk targeting. Instead, we focussed on a plausible age range and screening interval that closely approximates existing randomised screening evidence, albeit using PSA rather than MRI as the screening test. This allows for clearer analyses of strategies that are under consideration for prospective evaluation. Importantly, constraining the number of scenarios allows us to minimise, insofar as possible, hidden assumptions, such as relative mortality gains with much longer screening intervals. Alternative approaches to risk targeting and stratification will be the subject of further work.

## Conclusion

MRI-based screening for prostate cancer has the potential to reduce overdiagnoses and could generate more QALYs at an equivalent relative mortality reduction than PSA screening. However, strategies that triaged individuals prior to MRI, either through a polygenic risk score or PSA test, were both more cost-effective than MRI-first strategies and would be considered cost-effective within UK thresholds. Such strategies should be considered for further evaluation and potential implementation.

## Supporting information

Appendix B Results

## Data Availability

All data and code used in this analysis are publicly available from https://github.com/callta/mri-screening-prostate.

## Acknowledgements

During the course of this work, TC acknowledges funding from a Wellcome Clinical PhD Training Fellowship and an EU Horizon 2020 Grant (No 848294). NP is a co-investigator of the NIHR Policy Research Programme Unit on Cancer Awareness, Screening and Early Diagnosis (reference PR-PRU-NIHR206132)). The views expressed are those of the authors and not necessarily those of the NIHR or the Department of Health and Social Care.

## Contributor statement

TC: conceptualisation, project management, analysis, code development, writing – first draft and further edits. CM: writing – reviewing and editing. EF: writing – reviewing and editing. ME: writing – reviewing and editing. NP: conceptualisation, supervision, methodology, writing – review and editing. TC acts as guarantor.

## Ethics approval

This study was deemed exempt from formal ethical review because it only used unrestricted, publicly available, data.

