## Appendix A Methods for "The benefits, harms, and cost-effectiveness of age-based and risk-stratified screening for prostate cancer with MRI"

### Appendix Contents

**1. Model inputs** **2**

**2. Age-specific incidence and mortality data in the absence of screening** **5**

**3. Cancer incidence with screening** **5**

**4. Polygenic risk** **11**

**5. Utilities** **14**

**6. Resource use** **17**

**7. Costing descriptions** **17**

**References** **21**

### Appendix Tables

A.1 Model inputs and assumptions . . . . . . . . . . 3

A.2 Model parameters . . . . . . . . . . 4

A.3 Mean incidence and mortality rates from prostate cancer and other causes in England 2017–2019 6

A.4 Age-specific mean sojourn time . . . . . . . . . . 8

A.5 Age-specific percentage of screen-detected cancers overdiagnosed . . . . . . . . . . 10

A.6 Polygenic risk distribution . . . . . . . . . . 12

A.7 Age-specific stage at diagnosis and average utility values . . . . . . . . . . 15

A.8 Stage-specific utility values at 18 months from diagnosis . . . . . . . . . . 16

A.9 Resource use: Assessment of prostate cancers in base case scenarios . . . . . . . . . . 17

A.10 Detailed costings . . . . . . . . . . 18

### 1. Model inputs

**Appendix Table A.1:** Model inputs and assumptions

| Assumption | Description |
|---|---|
| Mean sojourn time (MST) | **MRI-first screening**:<br>• MST of 7.62 (95% intervals: 6.45–8.91) years.<br>• Cancers detected by screening considered clinically significant: 87%.<br><br>**PSA-first screening**:<br>• **Threshold 1ng/ml:** MST of 8.48 (95% intervals: 7.13–9.95) years.<br>• **Threshold 3ng/ml:** MST of 8.50 (95% intervals: 7.14–9.99) years.<br>• Cancers detected by screening considered clinically significant: 79%, irrespective of PSA threshold ($\geq$ 1ng/ml or $\geq$ 3ng/ml). We assumed reflex MRI testing after a positive PSA test would filter cancers by significance similarly between these two pools of potential cancers.<br><br>**Scenario analyses**: MST values between 7 and 10 years.<br>See subsection 3.1 for more details. |
| Relative mortality reduction with screening | • **MRI-first screening**: 0.82 (95% CI: 0.78–0.86)<br>• **PSA-first screening (1ng/ml)**: 0.83 (95% CI: 0.79–0.86)<br>• **PSA-first screening (3ng/ml)**: 0.87 (95% CI: 0.80–0.95) [1]<br><br>We do not assume that polygenic risk is associated with death from prostate cancer and so do not further adjust mortality rates applied.<br>**Scenario analyses**: Relative mortality reductions of between 0.7 and 0.9.<br>See subsection 3.3 for more details. |
| Episode sensitivity | The sensitivity is unknown for both an MRI-based and a PSA (1ng/ml followed by MRI with PI-RADS $\geq$ 4) screening episode. Using data from IP1-PROSTAGRAM [2] and the estimated sensitivity of multiparametric MRI within PSA screening [3, 4], we estimated the episode sensitivity as:<br>• **MRI-first screening**: 80% (95% CI: 69%–89%)<br>• **PSA-first screening (1ng/ml)**: 72% (95% CI: 62%–81%)<br>• **PSA-first screening (3ng/ml)**: 59% (95% CI: 49%–69%)<br><br>**Scenario analyses**: Episode sensitivity between 50% and 90% |

**Appendix Table A.2:** Model parameters

| Parameter | Value (95% CI) | Source |
|---|---|---|
| **Screening & diagnostic pathway** | | |
| *No screening* | | |
| Positive PSA test leading to a biopsy (clinically-suspected lesions) | 67% (55%–77%) | [5] |
| *MRI-first screening* | | |
| MRI screening tests positive (PI-RADS $\geq$ 4) | 10.6% (7.9%–14.0%) | [2] |
| *PSA-first screening* | | |
| PSA screening tests positive ($\geq$ 1ng/ml) | 35% (30%–40%) | [6, 7] |
| Positive PSA ($\geq$ 1ng/ml) and positive bpMRI (PI-RADS $\geq$ 4) | 7.5% (5.0%–10.0%) | [8] |
| **Costs, £ GBP** | | |
| Polygenic risk | 60 (28–105) | [a] |
| PSA test | 16 (7–28) | [9, 10] |
| Multiparametric MRI | 198 (91–345) | [11] |
| Biparametric MRI | 129 (60–225) | [11] |
| Transrectal ultrasound guided biopsy | 940 (434-1,638) | [11] |
| Staging costs | 567 (262–987) | [11] |
| Radical prostatectomy | 11,258 (5,196–19,611) | [11] |
| Radical radiotherapy | 7,749 (3,574–13,502) | [9, 10] |
| Chemotherapy | 2,586 (1,194–4,512) | [11–13] |
| Brachytherapy | 2,197 (1,014–3,830) | [9, 10] |
| Androgen deprivation therapy[d] | 804 (371–1,401) | [9, 10] |
| Active surveillance[b] | 6,366 (2,937–11,104) | [10, 14] |
| End of life costs | 11,074 (886–33,546) | [10, 15] |

[a] Personal communication of approximate cost to NHS.
[b] Total cost per cancer diagnosis (i.e. not yearly cost)
Abbreviations: MRI, magnetic resonance imaging; PSA, prostate specific antigen; CI, confidence intervals; GBP, British Pounds Sterling.

#### 2. Age-specific incidence and mortality data in the absence of screening

We used average 2017–2019 prostate cancer incidence and death data by 5-year age-group for those in England [16]. To translate rates by 5-year age-group into age-specific background rates (in the absence of screening), we fitted polynomial regression models of log rates against the midpoint age in each 5-year age-group. The final models used 3rd degree polynomials for prostate cancer incidence and mortality rates and a 2nd degree polynomial for mortality from other causes (Table A.3).

#### 3. Cancer incidence with screening

The natural history of prostate cancer involves a progression from an asymptomatic preclinical period to symptomatic clinical detection. The mean time between when a cancer could be detected by screening and when it would naturally progress to symptomatic presentation – is known as the mean sojourn time (MST).

The number of cancers on a prevalent (first) screen depends on the background pre-clinical incidence rate, $\lambda_1$, the sensitivity of a screening episode, $S$, and the pre-clinical screen-detectable period, the $MST$ [17]:

$$\text{Prevalent screen} = \lambda_1 \cdot S \cdot MST \tag{A.1}$$

The incidence rate at subsequent screens can be estimated using the following equation derived by Duffy et al. [18] from Day and Walter [19]:

$$\text{Incident screen} = \frac{pS\lambda_1(e^{-\lambda_1 t} - e^{-\lambda_2 t})}{(\lambda_2 - \lambda_1)(1 - (1 - S)e^{-\lambda_2 t})} \tag{A.2}$$

where screening uptake is denoted by $p$ and the screening interval by $t$. We use a simplifying assumption for both equations 1 and 2 that $\lambda_1$ can be approximated to the background clinical incidence of prostate cancer, $I$. Under the assumption that the transition hazard between preclinical and clinical disease follows an exponential distribution, the transition from pre-clinical to clinical (symptomatic) disease – the inverse of the MST – is denoted by $\lambda_2$.

Interval cancers at time, $t$, since the last screen depend on the age-specific background incidence of prostate cancer, screening sensitivity and the inverse of the MST ($\lambda_2$), and were estimated as [20]:

$$\text{Interval cancers } (t) = I(1 - S) + IS\Big(t + \frac{e^{-\lambda_2 t} - 1}{\lambda_2}\Big) \tag{A.3}$$

##### 3.1 Mean sojourn time

Using the age-specific values in Table A.4, we calculated a single age-specific weighted sum MST for each screening round based on the proportion expected to be diagnosed at different Gleason scores with screening. To account for uncertainty in these estimates, we modelled the MST from a Gamma distribution with a variance of 1.

**Appendix Table A.3:** Mean incidence and mortality rates from prostate cancer and other causes in England 2017–2019

| Age | Incidence rate | Mortality rate (prostate cancer) | Mortality rate (other causes) |
|---|---|---|---|
| 40 | 0.000017 | 0.000001 | 0.001465 |
| 41 | 0.000026 | 0.000001 | 0.001567 |
| 42 | 0.000038 | 0.000001 | 0.001678 |
| 43 | 0.000055 | 0.000002 | 0.001799 |
| 44 | 0.000079 | 0.000003 | 0.001931 |
| 45 | 0.000110 | 0.000003 | 0.002074 |
| 46 | 0.000151 | 0.000005 | 0.002231 |
| 47 | 0.000205 | 0.000006 | 0.002403 |
| 48 | 0.000274 | 0.000008 | 0.002590 |
| 49 | 0.000360 | 0.000010 | 0.002795 |
| 50 | 0.000466 | 0.000013 | 0.003020 |
| 51 | 0.000596 | 0.000017 | 0.003266 |
| 52 | 0.000751 | 0.000022 | 0.003536 |
| 53 | 0.000933 | 0.000028 | 0.003834 |
| 54 | 0.001145 | 0.000035 | 0.004160 |
| 55 | 0.001388 | 0.000044 | 0.004520 |
| 56 | 0.001662 | 0.000055 | 0.004916 |
| 57 | 0.001965 | 0.000068 | 0.005352 |
| 58 | 0.002298 | 0.000084 | 0.005834 |
| 59 | 0.002657 | 0.000103 | 0.006367 |
| 60 | 0.003039 | 0.000125 | 0.006956 |
| 61 | 0.003441 | 0.000152 | 0.007607 |
| 62 | 0.003856 | 0.000183 | 0.008329 |
| 63 | 0.004280 | 0.000219 | 0.009130 |
| 64 | 0.004706 | 0.000262 | 0.010019 |
| 65 | 0.005129 | 0.000311 | 0.011007 |
| 66 | 0.005542 | 0.000367 | 0.012105 |
| 67 | 0.005940 | 0.000433 | 0.013328 |
| 68 | 0.006316 | 0.000507 | 0.014691 |
| 69 | 0.006667 | 0.000592 | 0.016211 |
| 70 | 0.006987 | 0.000689 | 0.017908 |
| 71 | 0.007274 | 0.000799 | 0.019805 |
| 72 | 0.007525 | 0.000923 | 0.021928 |
| 73 | 0.007739 | 0.001063 | 0.024305 |
| 74 | 0.007914 | 0.001221 | 0.026969 |
| 75 | 0.008051 | 0.001397 | 0.029959 |
| 76 | 0.008151 | 0.001595 | 0.033318 |
| 77 | 0.008215 | 0.001816 | 0.037094 |
| 78 | 0.008246 | 0.002062 | 0.041344 |
| 79 | 0.008246 | 0.002337 | 0.046133 |
| 80 | 0.008219 | 0.002642 | 0.051533 |
| 81 | 0.008167 | 0.002982 | 0.057630 |
| 82 | 0.008095 | 0.003359 | 0.064520 |
| 83 | 0.008005 | 0.003778 | 0.072314 |
| 84 | 0.007901 | 0.004244 | 0.081140 |
| 85 | 0.007788 | 0.004761 | 0.091144 |
| 86 | 0.007667 | 0.005334 | 0.102496 |
| 87 | 0.007542 | 0.005972 | 0.115390 |
| 88 | 0.007417 | 0.006680 | 0.130050 |
| 89 | 0.007294 | 0.007468 | 0.146737 |

**MRI-first screening**: The proportion who might be diagnosed with a clinically significant cancer remains uncertain for a screening programme that used a screening MRI as the primary screening test. We used estimates from the Imperial Prostate 1 Prostate Cancer Screening Trial Using Imaging (IP1-PROSTAGRAM) [2] study and RE-IMAGINE [21] to anchor our assumptions. IP1-PROSTAGRAM involved an approximately 15 minutes-long non-contrast screening MRI. Amongst those who had a Prostate Imaging-Reporting and Data System (PIRADS)-2.1 score of 3-5, 14 of the 21 cancers detected on biopsy were considered clinically significant (67%, 95% confidence intervals [CI]: 45% to 83%). Restricting a positive scan to those with a PIRADS-2.1 score of 4-5 led to 69% (95% CI: 44% to 86%) of cancers detected being clinically significant. In RE-IMAGINE, 25 of 27 cancers detected in those with a positive MRI only were considered clinically significant (92.6%). A weighted average of these values gives 81%, which we subsequently multiplied by 1.07 (giving 87%) to take into account the relative proportion of clinically significant cancers diagnosed when using a PI-RADS threshold of 3 vs 4 in GÖTEBORG-2 [7]. This gave an estimated MST of 7.62 (95% intervals: 6.45–8.91) years.

**PSA-first screening**: In the European Randomized Study of Screening for Prostate Cancer (ERSPC) study, 33% of cancers were clinically significant [22], translating into an estimated MST of 12.63 (95% intervals: 11.16–14.24) years.

The proportion whose cancer might be detected at a clinically significant stage with a PSA-first screening strategy using a PSA threshold of 1ng/ml followed by an MRI with a PI-RADS score of $\geq 4$ is unclear. Both the GÖTEBORG-2 [7] and STHLM3-MRI studies [6] involved participants who received an MRI as a reflex test after screening positive with a PSA $\geq$3ng/ml. In the experimental arm of GÖTEBORG-2, multiparametric MRI was used as the reflex test; 62.5% of cancers detected were considered clinically significant [7]. In STHLM3-MRI, biparametric MRI was used; here, 82% of cancers detected were considered to be clinically significant [6]. Weighting based on the number of cancers detected, an average value for the percentage of cancers considered clinically significant for a PSA $\geq$ 3ng/ml & MRI with a PI-RADS $\geq$ 3 is 74%. We then multiplied this by 1.07 (giving 79%) to reflect the proportionate difference in those detected at a clinically significant stage when using a PI-RADS threshold of 4 vs 3 in GÖTEBORG-2. This translated into an MST of 8.48 (95% intervals: 7.13–9.95) years for PSA (threshold 1ng/ml) and an MST of 8.50 (95% intervals: 7.14–9.99) years for PSA (threshold 3ng/ml). The difference was due to the fact that PSA-first screening with a threshold of 1ng/ml was expected to have greater episode sensitivity, impacting the age-specific numbers screen-detected.

To account for the considerable uncertainty in these values, we run scenario analyses where the MST of screening strategies are explicitly varied.

##### 3.2 Life table specification

We used a life-table approach to model screening and no screening strategies. We initially used a population mortality rate, adjusted for the relative impact of screening where appropriate, to determine expected deaths amongst the screened and non-screened cohorts (see equations, algorithms, and code available at `https://github.com/callta/mri-screening-prostate` for more details).

**Appendix Table A.4:** Age-specific mean sojourn time

| Age group | Mean Sojourn Time (Gleason 6) | Mean Sojourn Time (Gleason $\geq$ 7) |
|---|---|---|
| 50-54 | 13.88 | 6.88 |
| 55-59 | 15.68 | 5.65 |
| 60-64 | 16.53 | 6.19 |
| 65-69 | 19.9 | 6.69 |
| 70-74 | 19.9 | 6.69 |

Age-specific values by Gleason score from the Cluster Randomized Trial of PSA Testing for Prostate Cancer (CAP) study [23, 24]

Using a population-based mortality rate allows us to model the population-level efficacy of prostate cancer screening amongst those who receive the intervention. In other words, we can use this to establish the numbers of cancer deaths in the screened arm under different relative mortality reduction with screening. But a population-level mortality rate doesn't allow us to disentangle deaths amongst those whose cancer was screen-detected or not. For a given screening interval and episode sensitivity, different mean sojourn times will mean that a different number of interval cancers could be expected between screening modalities (PSA or MRI). To better account for this, we calculated prostate cancer case hazards for cancers that are screen-detected and not screen detected.

We generated case hazards, taking into account lead time and the impact of different parameter combinations (e.g. mean sojourn time), by dividing the age-specific number of person-years of cancer by the number of prostate cancer deaths. Once we had calibrated this value for non-screen-detected cancers, we subsequently derived the case hazard for those whose cancer was screen-detected.

This allows us to determine the relative reduction in deaths specifically amongst those with a screen-detected cancers that is required to achieve a population-level efficacy (i.e. when considering both screen-detected and interval cancers, where the denominator is correspondingly greater). As an illustration, let us imagine that anyone diagnosed with cancer will die from their cancer and that there are 1,000 deaths from cancer in a cohort over a period of time. Were this cohort to undergo screening, we might expect only 800 deaths from cancer (i.e. a 20% relative reduction). The 200 deaths prevented must occur amongst those with a screen-detected cancer. So, if only 250 cancers were detected by screening, the relative mortality reduction amongst those who have a screen-detected cancer would be 80%.

##### 3.3 Mortality with screening

No data exist for an MRI-based screening programme. We assumed that the relative prostate-cancer-mortality reduction with an MRI-based screening programme could be at least that of a PSA-based programme [1]. In ERSPC, the maximal effect was 0.80 (95% CI: 0.67–0.93) [22, 25], reached after a run-in period of 10 years, with gradual waning as screening ends, approaching 0.87 (95% CI: 0.80–0.95) [1].

We applied the relative mortality rate from the ERPSC to PSA-first screening with a threshold of 3ng/ml. In our analyses, PSA-first screening involves a PSA test followed by reflex MRI. Because of this, the proportion of cancers expected to be detected at an insignificant stage will be lower than in the ERPSC trial where a positive PSA result led directly to a biopsy. Our results will thus be conservative.

We adjusted the relative mortality reductions with MRI-first and PSA-first screening (threshold 1ng/ml) to take account of different sensitivity and proportions of cancers detected at a clinically significant stage. We did this by applying the case hazards calculated for a PSA-first screening strategy with a threshold of 3ng/ml to the MRI-first and PSA-first (threshold 1ng/ml) strategies. This ensures the relative efficacy of screening, by which we mean the proportion of screen-detected cancers whose deaths were prevented by screening (as described in subsection 3.2 above), is approximately equal across the different strategies.

##### 3.4 Overdiagnosis

We derived the age-specific proportion of screen-detected cancers overdiagnosed by screening strategy using the lead time approach [17, 26, 27] (Table A.5) and applied this to the age-specific number of screen-detected cancers. We calculated the probability that a screen-detected cancers would be overdiagnosed as $e^{-\lambda t}$, where $t$ is remaining life-expectancy and $\lambda = 1/MST$ [17]. The lead time approach assumes that a cancer would have gone on to be clinically diagnosed if the individual had lived long enough, such that the numbers shown to be overdiagnosed in this analysis can reasonably be considered as a lower estimate.

We present estimates using the lead time approach to set a minimum floor on the percentage of screen-detected cancers likely to be overdiagnosed. This is because, with uptake and compliance set to 100% coupled with strategies where screening is restricted to those at very high risk, overdiagnosis can become negative with the excess incidence approach. In other words, such screening strategies might lead to fewer overdiagnosed than no screening. In theory, it could be possible that a risk-stratified programme restricted to those at very high risk with perfect adherence and uptake might reduce overdiagnosis relative to no screening. This is because currently ad hoc screening occurs that will inevitably cause overdiagnosis that we are not otherwise accounting for. To calculate overdiagnosis using excess cumulative incidence from our tables, subtract the incident prostate cancers by screening strategy from those that occur with no screening.

**Appendix Table A.5:** Age-specific percentage of screen-detected cancers overdiagnosed

| Age (years) | PSA $\geq$ 1 ng/ml | PSA $\geq$ 3 ng/ml | MRI |
|---|---|---|---|
| 50 | 6.22 | 6.22 | 5.52 |
| 51 | 6.68 | 6.68 | 5.94 |
| 52 | 7.17 | 7.17 | 6.39 |
| 53 | 7.71 | 7.71 | 6.88 |
| 54 | 8.28 | 8.28 | 7.40 |
| 55 | 7.91 | 7.91 | 6.67 |
| 56 | 8.52 | 8.52 | 7.20 |
| 57 | 9.16 | 9.16 | 7.77 |
| 58 | 9.86 | 9.86 | 8.38 |
| 59 | 10.6 | 10.6 | 9.04 |
| 60 | 12.67 | 12.67 | 10.94 |
| 61 | 13.58 | 13.58 | 11.76 |
| 62 | 14.56 | 14.56 | 12.65 |
| 63 | 15.59 | 15.59 | 13.60 |
| 64 | 16.69 | 16.69 | 14.61 |
| 65 | 20.72 | 20.72 | 17.97 |
| 66 | 22.05 | 22.05 | 19.21 |
| 67 | 23.44 | 23.44 | 20.52 |
| 68 | 24.91 | 24.91 | 21.90 |
| 69 | 26.45 | 26.45 | 23.36 |
| 70 | 28.06 | 28.06 | 24.90 |
| 71 | 29.74 | 29.74 | 26.51 |
| 72 | 31.49 | 31.49 | 28.20 |
| 73 | 33.32 | 33.32 | 29.97 |
| 74 | 35.21 | 35.21 | 31.82 |
| 75 | 37.17 | 37.17 | 33.74 |
| 76 | 39.20 | 39.20 | 35.74 |
| 77 | 41.29 | 41.29 | 37.81 |
| 78 | 43.44 | 43.44 | 39.95 |
| 79 | 45.64 | 45.64 | 42.16 |

Average age-specific values by screening modality (PSA using a threshold of 1ng/ml or 3ng/ml, or MRI) for base-case analyses. For example, 6.67% of cancers that were detected through screening with MRI at age 55 could be expected to be overdiagnosed. The percentage of screen-detected cancers overdiagnosed varies by the MST. Correspondingly, these values were recalculated in relevant scenario analyses in which the MST was varied.

### 4. Polygenic risk

In Figure A.1 and Table A.6, we show the proportion of cases and the relative risks of prostate cancer above and below each centile of the polygenic risk distribution. In risk-based screening analyses, we apply the relative risks to incidence rates in relevant cohorts. Age-specific 10-year and lifetime absolute risks of prostate cancer were calculated using Devcan software [28].

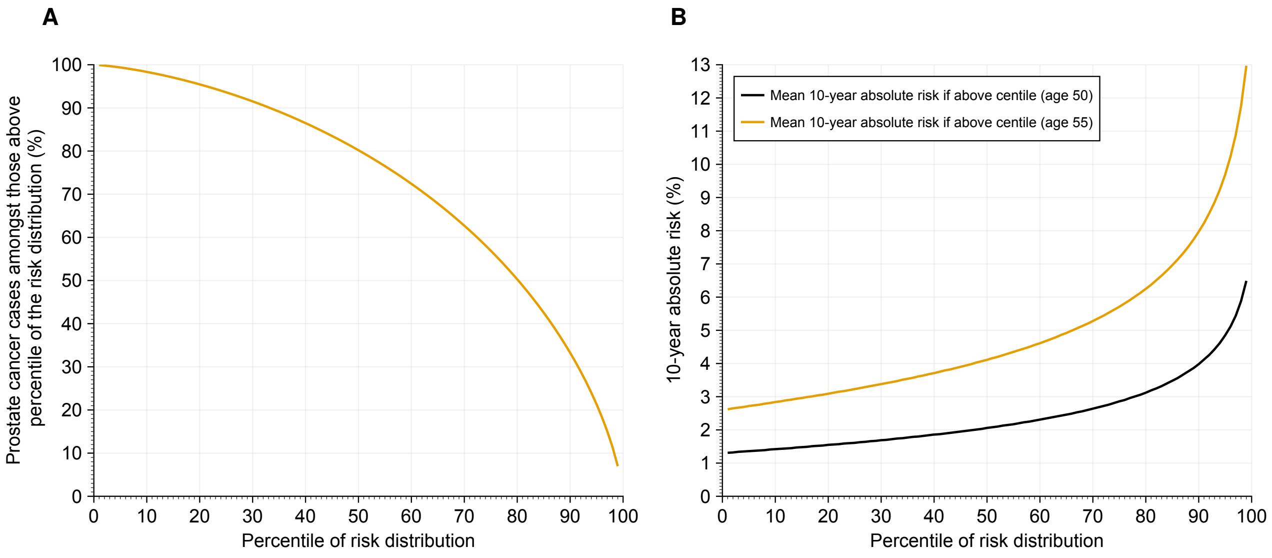

**Appendix Figure A.1:** Subfigure A shows the percentage of prostate cancers expected amongst those above each centile of the polygenic risk distribution. For example, 80% of prostate cancers are expected to occur amongst those above the 50th percentile of the risk distribution. Subfigure B shows the mean 10-year absolute risk of prostate cancer by percentile of the risk distribution from the age of 50 (black) and 55 (blue). The average 10-year risk of prostate cancer doubles from 1.3% to 2.6% between the age of 50 and 55, but lifetime risks remain approximately equal at 15%. Correspondingly, a risk-based screening programme from age 55 that uses a 3.5% 10-year absolute risk threshold is screening those from the approximately 35th centile (subfigure B) and so could be expected to include approximately 90% of men who will go on to develop prostate cancer in their lifetime (subfigure A). The same 10-year absolute risk threshold applied to a screening programme starting at age 50 involves screening those men at the approximately 85th centile (subfigure B), and would include 43% of men who will go on to develop prostate cancer in their lifetime (subfigure A).

**Appendix Table A.6:** Polygenic risk distribution

| Percentile of risk distribution | Proportion above centile (%) | Cases amongst those above centile (%) | RR of PCa diagnosis if above centile | RR of Pca diagnosis if below centile | 10-year absolute risk at age 55 if above centile (%) | 10-year absolute risk of PCa at age 55 if below centile (%) | Lifetime absolute risk of PCa at age 55 if above centile (%) | Lifetime absolute risk of PCa at age 55 if below centile (%) |
|---|---|---|---|---|---|---|---|---|
| 1 | 99 | 99.93 | 1.01 | 0.10 | 2.62 | 0.26 | 15.34 | 1.54 |
| 2 | 98 | 99.81 | 1.02 | 0.11 | 2.65 | 0.30 | 15.47 | 1.74 |
| 3 | 97 | 99.68 | 1.03 | 0.13 | 2.67 | 0.33 | 15.61 | 1.91 |
| 4 | 96 | 99.53 | 1.04 | 0.14 | 2.69 | 0.35 | 15.75 | 2.06 |
| 5 | 95 | 99.37 | 1.05 | 0.14 | 2.72 | 0.38 | 15.88 | 2.19 |
| 6 | 94 | 99.19 | 1.05 | 0.15 | 2.74 | 0.40 | 16.02 | 2.32 |
| 7 | 93 | 98.99 | 1.06 | 0.16 | 2.76 | 0.42 | 16.16 | 2.44 |
| 8 | 92 | 98.79 | 1.07 | 0.17 | 2.79 | 0.44 | 16.3 | 2.55 |
| 9 | 91 | 98.57 | 1.08 | 0.18 | 2.81 | 0.46 | 16.44 | 2.66 |
| 10 | 90 | 98.34 | 1.09 | 0.18 | 2.84 | 0.47 | 16.58 | 2.77 |
| 11 | 89 | 98.10 | 1.1 | 0.19 | 2.86 | 0.49 | 16.72 | 2.87 |
| 12 | 88 | 97.85 | 1.11 | 0.20 | 2.89 | 0.51 | 16.87 | 2.97 |
| 13 | 87 | 97.59 | 1.12 | 0.20 | 2.91 | 0.53 | 17.01 | 3.07 |
| 14 | 86 | 97.31 | 1.13 | 0.21 | 2.94 | 0.54 | 17.16 | 3.17 |
| 15 | 85 | 97.03 | 1.14 | 0.21 | 2.96 | 0.56 | 17.31 | 3.26 |
| 16 | 84 | 96.73 | 1.15 | 0.22 | 2.99 | 0.57 | 17.46 | 3.36 |
| 17 | 83 | 96.43 | 1.16 | 0.23 | 3.01 | 0.59 | 17.61 | 3.45 |
| 18 | 82 | 96.11 | 1.17 | 0.23 | 3.04 | 0.61 | 17.76 | 3.54 |
| 19 | 81 | 95.79 | 1.18 | 0.24 | 3.06 | 0.62 | 17.91 | 3.63 |
| 20 | 80 | 95.45 | 1.19 | 0.24 | 3.09 | 0.64 | 18.07 | 3.72 |
| 21 | 79 | 95.10 | 1.20 | 0.25 | 3.12 | 0.65 | 18.23 | 3.80 |
| 22 | 78 | 94.75 | 1.21 | 0.26 | 3.15 | 0.67 | 18.39 | 3.89 |
| 23 | 77 | 94.38 | 1.22 | 0.26 | 3.17 | 0.68 | 18.55 | 3.98 |
| 24 | 76 | 94.00 | 1.23 | 0.27 | 3.20 | 0.70 | 18.72 | 4.06 |
| 25 | 75 | 93.61 | 1.24 | 0.27 | 3.23 | 0.71 | 18.88 | 4.15 |
| 26 | 74 | 93.21 | 1.25 | 0.28 | 3.26 | 0.72 | 19.05 | 4.24 |
| 27 | 73 | 92.80 | 1.26 | 0.28 | 3.29 | 0.74 | 19.22 | 4.32 |
| 28 | 72 | 92.38 | 1.28 | 0.29 | 3.32 | 0.75 | 19.40 | 4.41 |
| 29 | 71 | 91.95 | 1.29 | 0.30 | 3.35 | 0.77 | 19.57 | 4.49 |
| 30 | 70 | 91.51 | 1.30 | 0.30 | 3.38 | 0.78 | 19.75 | 4.58 |
| 31 | 69 | 91.06 | 1.31 | 0.31 | 3.41 | 0.80 | 19.93 | 4.66 |
| 32 | 68 | 90.60 | 1.32 | 0.31 | 3.44 | 0.81 | 20.12 | 4.75 |
| 33 | 67 | 90.12 | 1.34 | 0.32 | 3.47 | 0.83 | 20.30 | 4.84 |
| 34 | 66 | 89.63 | 1.35 | 0.32 | 3.51 | 0.84 | 20.49 | 4.92 |
| 35 | 65 | 89.14 | 1.36 | 0.33 | 3.54 | 0.86 | 20.69 | 5.01 |
| 36 | 64 | 88.63 | 1.37 | 0.34 | 3.57 | 0.87 | 20.88 | 5.09 |
| 37 | 63 | 88.11 | 1.39 | 0.34 | 3.61 | 0.89 | 21.08 | 5.18 |
| 38 | 62 | 87.58 | 1.40 | 0.35 | 3.64 | 0.90 | 21.29 | 5.27 |
| 39 | 61 | 87.03 | 1.41 | 0.35 | 3.68 | 0.92 | 21.49 | 5.36 |
| 40 | 60 | 86.47 | 1.43 | 0.36 | 3.71 | 0.93 | 21.70 | 5.44 |
| 41 | 59 | 85.91 | 1.44 | 0.36 | 3.75 | 0.95 | 21.92 | 5.53 |
| 42 | 58 | 85.32 | 1.46 | 0.37 | 3.79 | 0.96 | 22.14 | 5.62 |
| 43 | 57 | 84.73 | 1.47 | 0.38 | 3.82 | 0.98 | 22.36 | 5.71 |
| 44 | 56 | 84.12 | 1.49 | 0.38 | 3.86 | 0.99 | 22.59 | 5.80 |
| 45 | 55 | 83.50 | 1.50 | 0.39 | 3.90 | 1.01 | 22.82 | 5.89 |
| 46 | 54 | 82.87 | 1.52 | 0.39 | 3.94 | 1.02 | 23.05 | 5.98 |
| 47 | 53 | 82.22 | 1.53 | 0.40 | 3.98 | 1.04 | 23.29 | 6.07 |
| 48 | 52 | 81.56 | 1.55 | 0.41 | 4.03 | 1.05 | 23.54 | 6.16 |
| 49 | 51 | 80.88 | 1.57 | 0.41 | 4.07 | 1.07 | 23.79 | 6.26 |
| 50 | 50 | 80.19 | 1.58 | 0.42 | 4.11 | 1.09 | 24.05 | 6.35 |
| 51 | 49 | 79.49 | 1.60 | 0.42 | 4.16 | 1.10 | 24.31 | 6.45 |
| 52 | 48 | 78.77 | 1.62 | 0.43 | 4.20 | 1.12 | 24.58 | 6.54 |

*Continued on the next page*

**Appendix Table A.6**: Polygenic risk distribution, continued

| Percentile of risk distribution | Proportion above centile (%) | Cases amongst those above centile (%) | RR of PCa diagnosis if above centile | RR of Pca diagnosis if below centile | 10-year absolute risk at age 55 if above centile (%) | 10-year absolute risk of PCa at age 55 if below centile (%) | Lifetime absolute risk of PCa at age 55 if above centile (%) | Lifetime absolute risk of PCa at age 55 if below centile (%) |
|---|---|---|---|---|---|---|---|---|
| 53 | 47 | 78.03 | 1.64 | 0.44 | 4.25 | 1.14 | 24.85 | 6.64 |
| 54 | 46 | 77.28 | 1.65 | 0.44 | 4.30 | 1.15 | 25.13 | 6.74 |
| 55 | 45 | 76.51 | 1.67 | 0.45 | 4.35 | 1.17 | 25.42 | 6.84 |
| 56 | 44 | 75.73 | 1.69 | 0.46 | 4.40 | 1.19 | 25.72 | 6.94 |
| 57 | 43 | 74.93 | 1.71 | 0.46 | 4.45 | 1.20 | 26.02 | 7.04 |
| 58 | 42 | 74.11 | 1.73 | 0.47 | 4.50 | 1.22 | 26.33 | 7.14 |
| 59 | 41 | 73.27 | 1.75 | 0.48 | 4.56 | 1.24 | 26.65 | 7.24 |
| 60 | 40 | 72.41 | 1.77 | 0.48 | 4.61 | 1.26 | 26.98 | 7.35 |
| 61 | 39 | 71.54 | 1.80 | 0.49 | 4.67 | 1.28 | 27.31 | 7.45 |
| 62 | 38 | 70.65 | 1.82 | 0.50 | 4.73 | 1.29 | 27.66 | 7.56 |
| 63 | 37 | 69.73 | 1.84 | 0.50 | 4.79 | 1.31 | 28.02 | 7.67 |
| 64 | 36 | 68.80 | 1.87 | 0.51 | 4.86 | 1.33 | 28.39 | 7.78 |
| 65 | 35 | 67.84 | 1.89 | 0.52 | 4.92 | 1.35 | 28.77 | 7.90 |
| 66 | 34 | 66.86 | 1.92 | 0.53 | 4.99 | 1.37 | 29.16 | 8.01 |
| 67 | 33 | 65.86 | 1.94 | 0.53 | 5.06 | 1.39 | 29.56 | 8.13 |
| 68 | 32 | 64.83 | 1.97 | 0.54 | 5.13 | 1.41 | 29.98 | 8.24 |
| 69 | 31 | 63.78 | 2.00 | 0.55 | 5.20 | 1.43 | 30.41 | 8.37 |
| 70 | 30 | 62.71 | 2.03 | 0.56 | 5.28 | 1.45 | 30.86 | 8.49 |
| 71 | 29 | 61.61 | 2.06 | 0.57 | 5.36 | 1.47 | 31.33 | 8.61 |
| 72 | 28 | 60.48 | 2.09 | 0.58 | 5.44 | 1.50 | 31.81 | 8.74 |
| 73 | 27 | 59.32 | 2.13 | 0.58 | 5.53 | 1.52 | 32.31 | 8.87 |
| 74 | 26 | 58.13 | 2.16 | 0.59 | 5.62 | 1.54 | 32.84 | 9.00 |
| 75 | 25 | 56.91 | 2.20 | 0.60 | 5.71 | 1.56 | 33.38 | 9.14 |
| 76 | 24 | 55.65 | 2.23 | 0.61 | 5.81 | 1.59 | 33.95 | 9.28 |
| 77 | 23 | 54.37 | 2.27 | 0.62 | 5.91 | 1.61 | 34.55 | 9.42 |
| 78 | 22 | 53.04 | 2.31 | 0.63 | 6.02 | 1.64 | 35.18 | 9.57 |
| 79 | 21 | 51.68 | 2.36 | 0.64 | 6.13 | 1.66 | 35.84 | 9.71 |
| 80 | 20 | 50.28 | 2.40 | 0.65 | 6.25 | 1.69 | 36.53 | 9.87 |
| 81 | 19 | 48.83 | 2.45 | 0.66 | 6.37 | 1.71 | 37.26 | 10.03 |
| 82 | 18 | 47.34 | 2.50 | 0.67 | 6.51 | 1.74 | 38.03 | 10.19 |
| 83 | 17 | 45.79 | 2.56 | 0.68 | 6.65 | 1.77 | 38.85 | 10.36 |
| 84 | 16 | 44.20 | 2.61 | 0.69 | 6.80 | 1.80 | 39.73 | 10.53 |
| 85 | 15 | 42.55 | 2.68 | 0.70 | 6.96 | 1.83 | 40.66 | 10.71 |
| 86 | 14 | 40.84 | 2.74 | 0.72 | 7.13 | 1.86 | 41.66 | 10.89 |
| 87 | 13 | 39.06 | 2.81 | 0.73 | 7.31 | 1.90 | 42.74 | 11.08 |
| 88 | 12 | 37.20 | 2.89 | 0.74 | 7.51 | 1.93 | 43.91 | 11.28 |
| 89 | 11 | 35.27 | 2.97 | 0.76 | 7.73 | 1.97 | 45.18 | 11.49 |
| 90 | 10 | 33.25 | 3.06 | 0.77 | 7.97 | 2.00 | 46.58 | 11.71 |
| 91 | 9 | 31.13 | 3.17 | 0.79 | 8.23 | 2.04 | 48.13 | 11.94 |
| 92 | 8 | 28.89 | 3.28 | 0.80 | 8.53 | 2.08 | 49.85 | 12.19 |
| 93 | 7 | 26.52 | 3.41 | 0.82 | 8.86 | 2.13 | 51.80 | 12.44 |
| 94 | 6 | 24.00 | 3.56 | 0.84 | 9.24 | 2.18 | 54.04 | 12.72 |
| 95 | 5 | 21.29 | 3.73 | 0.86 | 9.69 | 2.23 | 56.65 | 13.02 |
| 96 | 4 | 18.35 | 3.93 | 0.88 | 10.23 | 2.28 | 59.78 | 13.34 |
| 97 | 3 | 15.10 | 4.19 | 0.90 | 10.89 | 2.34 | 63.66 | 13.70 |
| 98 | 2 | 11.41 | 4.52 | 0.93 | 11.75 | 2.41 | 68.72 | 14.11 |
| 99 | 1 | 6.97 | 4.99 | 0.96 | 12.97 | 2.50 | 75.84 | 14.59 |

Polygenic risk distribution given a variance of 0.72. At age 55, the average 10-year risk is 2.6% and the average lifetime risk is 15.2%.

Abbreviations: RR, relative risk. PCa, prostate cancer.

#### 5. Utilities

We used age-stratified EQ-5D-5L scores for the general population from the 2018 Health Survey for England [29]. As these were available in five-yearly age groups, we used polynomial regression (with age as a second-degree polynomial) to interpolate yearly values (Appendix Table A.7). We calculated an average utility values for those with prostate cancer, considering:

- The age-specific distribution of cancer stage at diagnosis (average 2017-2019 from NHS Digital [30]);
- the proportions receiving active surveillance or radical therapy by stage [31]; and
- the expected median survival based on age at diagnosis [32].

We considered radical therapy and active surveillance to have utilities of 0.78 [33] and 0.88, respectively. We assume 55% of those having active surveillance transition to radical therapy, as in the ProtecT study.

The 2024 NHS Cancer Quality of Life Survey provides stage-specific EQ-5D-5L scores of individuals with prostate cancer 18 months after their diagnosis (Appendix Table A.8) [34]. This survey has been completed by approximately 50% of all individuals with a prostate cancer diagnosed in England since autumn 2021.

We assumed that a diagnosis of prostate cancer would only attract a lower utility than the age-specific average background value for ten years. Tan and colleagues, using comprehensive data from the UK Clinical Practice Data Link (CPRD), estimated median survival after a prostate cancer diagnosis of 11 years [32]. Unsurprisingly, there were significant differences by age at diagnosis, with median survival of approximately 18 years if diagnosed between the ages of 60-69, 10 years if diagnosed between 70-79, and <5 years if diagnosed from 80 onwards [32].

**Appendix Table A.7:** Age-specific stage at diagnosis and average utility values

| | Stage at diagnosis (%) | | | | | |
|---|---|---|---|---|---|---|
| Age | 1 | 2 | 3 | 4 | Background utility | Utility with prostate cancer |
| 50 | 0.324 | 0.398 | 0.128 | 0.150 | 0.825 | 0.808 |
| 51 | 0.324 | 0.398 | 0.128 | 0.150 | 0.822 | 0.805 |
| 52 | 0.324 | 0.398 | 0.128 | 0.150 | 0.819 | 0.802 |
| 53 | 0.324 | 0.398 | 0.128 | 0.150 | 0.816 | 0.799 |
| 54 | 0.324 | 0.398 | 0.128 | 0.150 | 0.814 | 0.797 |
| 55 | 0.303 | 0.377 | 0.141 | 0.179 | 0.812 | 0.794 |
| 56 | 0.303 | 0.377 | 0.141 | 0.179 | 0.811 | 0.792 |
| 57 | 0.303 | 0.377 | 0.141 | 0.179 | 0.809 | 0.791 |
| 58 | 0.303 | 0.377 | 0.141 | 0.179 | 0.808 | 0.790 |
| 59 | 0.303 | 0.377 | 0.141 | 0.179 | 0.808 | 0.790 |
| 60 | 0.266 | 0.353 | 0.164 | 0.217 | 0.807 | 0.787 |
| 61 | 0.266 | 0.353 | 0.164 | 0.217 | 0.807 | 0.787 |
| 62 | 0.266 | 0.353 | 0.164 | 0.217 | 0.806 | 0.786 |
| 63 | 0.266 | 0.353 | 0.164 | 0.217 | 0.806 | 0.786 |
| 64 | 0.266 | 0.353 | 0.164 | 0.217 | 0.806 | 0.786 |
| 65 | 0.239 | 0.343 | 0.174 | 0.244 | 0.806 | 0.784 |
| 66 | 0.239 | 0.343 | 0.174 | 0.244 | 0.805 | 0.784 |
| 67 | 0.239 | 0.343 | 0.174 | 0.244 | 0.805 | 0.784 |
| 68 | 0.239 | 0.343 | 0.174 | 0.244 | 0.805 | 0.783 |
| 69 | 0.239 | 0.343 | 0.174 | 0.244 | 0.804 | 0.783 |
| 70 | 0.187 | 0.313 | 0.193 | 0.307 | 0.803 | 0.760 |
| 71 | 0.187 | 0.313 | 0.193 | 0.307 | 0.802 | 0.759 |
| 72 | 0.187 | 0.313 | 0.193 | 0.307 | 0.801 | 0.758 |
| 73 | 0.187 | 0.313 | 0.193 | 0.307 | 0.800 | 0.756 |
| 74 | 0.187 | 0.313 | 0.193 | 0.307 | 0.798 | 0.755 |
| 75 | 0.161 | 0.293 | 0.195 | 0.351 | 0.796 | 0.749 |
| 76 | 0.161 | 0.293 | 0.195 | 0.351 | 0.793 | 0.746 |
| 77 | 0.161 | 0.293 | 0.195 | 0.351 | 0.790 | 0.743 |
| 78 | 0.161 | 0.293 | 0.195 | 0.351 | 0.786 | 0.740 |
| 79 | 0.161 | 0.293 | 0.195 | 0.351 | 0.782 | 0.736 |
| 80 | 0.127 | 0.235 | 0.192 | 0.445 | 0.778 | 0.712 |
| 81 | 0.127 | 0.235 | 0.192 | 0.445 | 0.773 | 0.707 |
| 82 | 0.127 | 0.235 | 0.192 | 0.445 | 0.767 | 0.702 |
| 83 | 0.127 | 0.235 | 0.192 | 0.445 | 0.760 | 0.696 |
| 84 | 0.127 | 0.235 | 0.192 | 0.445 | 0.753 | 0.689 |
| 85 | 0.124 | 0.184 | 0.164 | 0.528 | 0.746 | 0.676 |
| 86 | 0.124 | 0.184 | 0.164 | 0.528 | 0.737 | 0.668 |
| 87 | 0.124 | 0.184 | 0.164 | 0.528 | 0.727 | 0.660 |
| 88 | 0.124 | 0.184 | 0.164 | 0.528 | 0.717 | 0.651 |
| 89 | 0.124 | 0.184 | 0.164 | 0.528 | 0.706 | 0.640 |

**Appendix Table A.8:** Stage-specific utility values at 18 months from diagnosis

| Stage | EQ-5D score | Relative decrement[a] |
|---|---|---|
| 1 | 80.45 | 1 |
| 2 | 80.43 | 1 |
| 3 | 77.68 | 0.98 |
| 4 | 69.59 | 0.88 |

[a] Decrement relative to average background EQ-5D score from ages 50-89 (0.791) as shown in Appendix Table A.7.
Results are from the NHS England Cancer Quality of Life Survey, March 2024 [34].

### 6. Resource use

**Appendix Table A.9:** Resource use: Assessment of prostate cancers in base case scenarios

| Strategy | Description |
|---|---|
| Non-screened cohort | Based on previous work [14], we used an estimate of 1.88 mpMRI scans required to detect one cancer, using results from the PROMIS trial [35]. Multiparametric MRI is the current recommended diagnostic standard for those with a positive PSA and clinically suspected prostate cancer in the UK [36]. We assumed that there were 20% more PSA tests than MRI scans [9]. We estimated that two-thirds of those having an mpMRI would go on to have a biopsy (67%, 95% CI: 55%-77%) [5]. |
| MRI-first screening | **PI-RADS $\geq$ 4**: 10.6% (7.9%–14.0%): We used the estimates of the percentage of those with a positive MRI from IP1-PROSTAGRAM [2][a]. |
| PSA-first screening[b] | **PSA $\geq$ 1ng/ml & PI-RADS $\geq$ 4**: We estimated the proportion of PSA tests that would screen positive with a threshold of 1ng/ml as 35%. This was based on the proportion of those in STHLM-3 [6] and GÖTEBORG-2 [7] with PSA values greater than 1.5ng/ml and 1.2ng/ml, respectively. We estimated 7.5% (95% CI: 5.0%–10%) would have both a positive PSA ($\geq$1ng/ml) and MRI (PI-RADS score $\geq$ 4) based on IP1-PROSTAGRAM [8].<br><br>**PSA $\geq$ 3ng/ml & PI-RADS $\geq$ 4**: 16.8% (16.6%–17.0%) of PSA tests returned a result of $\geq$ 3ng/ml in ERSPC [25]. We estimated 18% of those with a PSA $\geq$ 3ng/ml would also have a MRI with a PI-RADS score of $\geq$ 4, based on STHLM-3 [6]. |

[a] Test positivity would be expected to reduce in subsequent rounds of screening as the number of screen-detected cancers reduces after the prevalence screen. However, in the absence of follow-on data from further screening rounds, we have used the prevalence scan estimates throughout and vary this value in scenario analyses.

[b] The PRIME trial has shown that bpMRI is not inferior to mpMRI for the diagnosis of clinically significant prostate cancer [37]. We therefore considered that a PSA screening programme, were one to be proposed today, would likely use bpMRI as a reflex test. This is congruent with the TRANSFORM trial where bpMRI is being used as a primary screening test with or without PSA [38].

### 7. Costing descriptions

We included costs for screening, assessment, diagnosis, treatment, and palliation and death from prostate cancer. Pathways are based on NICE guidelines [36]. Individual resource costs are presented in Appendix Table A.2 and A.10. Given uncertainties in their estimation, we modelled costs using a Gamma distribution where the spread was the central estimate +/- 1/3 and subject them to probabilistic one-way sensitivity analyses. All costs use either 2025/2026 NHS Tariffs or have been inflated to prices as of August 2025.

**Appendix Table A.10:** Detailed costings

| Resource | Description | Cost (£) | Source |
|---|---|---|---|
| PSA test | Cost of phlebotomy and biochemistry | 16 (7–28) | [9, 10] |
| Multiparametric (mp)-MRI | NHS 2025/26 tariff cost (RD03Z) | 198 (91–345) | [11] |
| Biparametric (bp)-MRI | The cost of a non-contrast MRI scan on one area is £129 (code RD01A) including reading the scan. This tallies with the cost considered in the PRIME trial for a biparametric MRI [37]. | 129 (60–225) | [11] |
| Transrectal ultrasound guided biopsy | NHS 2025/26 tariff code LB76Z for a transrectal ultrasound guided biopsy of the prostate (£491) combined with one urological appointment (code 101 – WF01B first attendance, single professional in a Urology Service, £164) and the cost of histopathology (£185 after inflation to 2025 values [39]). 1.3% (95% CI: 0.8%–2.1%) were estimated to be admitted after a biopsy with sepsis (ProtecT study [40]). The mean cost of a non-elective spell in hospital with sepsis (NHS Tariff Codes WJ06A-J: Sepsis with & without interventions) was rounded to £7,682. Total cost: £491 + £164 + 185 + (0.013 $\times$ £7682) | 940 (434–1,637) | [11] |
| Staging costs | We considered the following costs for staging a prostate cancer:<br>• Urological appointment £107 Source: NHS 2025/26 tariff, code: 101<br>• Isotope bone scan £249 including reporting source: NHS 2025/26 tariff, code: RN15A<br>• Urology MDT £211 Source: NHS 2025/26 tariff, code: 101 multi-professional (WF02B)<br>Not all patients will receive a bone scan, and some may have a CT pelvis. We account for this uncertainty probabilistically. | 567 (262–987) | [11] |

*Continued on next page*

**Appendix Table A.10**: Costings (August 2025), continued

| Resource | Description | Cost (£) | Source |
|---|---|---|---|
| Radical prostatectomy | Urological appointment £107 Source: NHS 2025/26 tariff, code: 101 In 2019, the most recent (pre-COVID) year for which the National Prostate Cancer Audit has reported, 89% of prostatectomies were robotic, 6% laproscopic, and 5% open. The guide prices for these are £9,186, £8,277, and £8,804 (codes LB69Z, LB22Z and mean of £11,131 and £6,476 for LB21A/B). In addition, 14% were re-admitted as an emergency within 90-days of radical prostatectomy. We have not estimated the impact of complications such as incontinence directly. | 11,258 (5,196–19,611) | [11] |
| Radical radiotherapy | Appointment with a clinical oncologist, preparation for IMRT, its delivery and adverse effects. From Callender et al. 2019 [9] with uplift for inflation. | 7,749 (3,574–13,502) | [9, 10] |
| Chemotherapy | Assumptions follow NICE resource impact assessment:<br>• First (£322) outpatient and one follow-up appointment (£150) with a clinical oncologist (code 800 of NHS tariff 2025/26).<br>• Cost of delivery of simple parenteral chemotherapy at the first attendance (£182) (SB12Z), followed by the cost of subsequent rounds of chemotherapy (SB15Z) (£265).<br>• Up to 10 cycles of docetaxel are recommended, however an average of 6 has been used based on the NICE guideline resource impact report.<br>• Docetaxel costs £17.88 for 160mg/8ml solution, so average cost of $17.88 \times 6$ = £107.28 | 2,586 (1,194–4,512) | [11–13] |
| Brachytherapy | Sourced from Callender et al. 2019, inflated to 2025 costs. | 2,197 (1,014–3,830) | [9, 10] |

*Continued on next page*

**Appendix Table A.10**: Costings (August 2025), continued

| Resource | Description | Cost (£) | Source |
|---|---|---|---|
| Androgen deprivation therapy | Sourced from Callender et al. 2019, inflated to 2025 costs. | 804 (371–1,401) | [9, 10] |
| Active surveillance | Sourced from Callender et al. 2021, inflated to 2025 costs. | 6,366 (2937–11,104) | [10, 14] |
| End of life costs | Sourced from Round and colleagues, inflated to 2025 values | 11,074 (886–33,546) | [10, 15] |
