## Appendix B Results for "The benefits, harms, and cost-effectiveness of age-based and risk-stratified screening for prostate cancer with MRI"

### Appendix B Figures


B.1 ERSPC comparison . . . . . . . . . . 2
B.2 Cumulative incidence, deaths from cancer, and deaths from other causes with MRI screening . 3
B.3 Scenario analysis at a WTP of £35,000 per QALY (MRI-first screening) . . . . . . . . . . 4
B.4 NHB by WTP threshold (MRI-first screening, PI-RADS $\geq$ 3) . . . . . . . . . . 5
B.5 Probabilistic one-way sensitivity analysis (MRI-first screening) . . . . . . . . . . 6
B.6 Mortality: all screened and screen-detected . . . . . . . . . . 7


### Appendix B Tables


B.1 Outcomes per 1,000 men with risk-stratified MRI-first screening . . . . . . . . . . 8
B.2 Overdiagnosis estimated using excess incidence and lead time methods . . . . . . . . . . 11
B.3 Screening strategies ranked by the probability that their incremental net health benefit exceeds zero at £25,000 per QALY . . . . . . . . . . 12
B.4 Scenario analysis: The impact of varying the relative impact of screening on prostate cancer-specific mortality . . . . . . . . . . 14
B.5 Scenario analysis: The impact of varying the mean sojourn time (MST) on screening . . . . . 17
B.6 Scenario analysis: The impact of varying the age at which screening starts and stops . . . . . . 19
B.7 Outcomes per 1,000 men for age-based and risk-based MRI-first screening (PI-RADS $\geq$ 3) . . 22

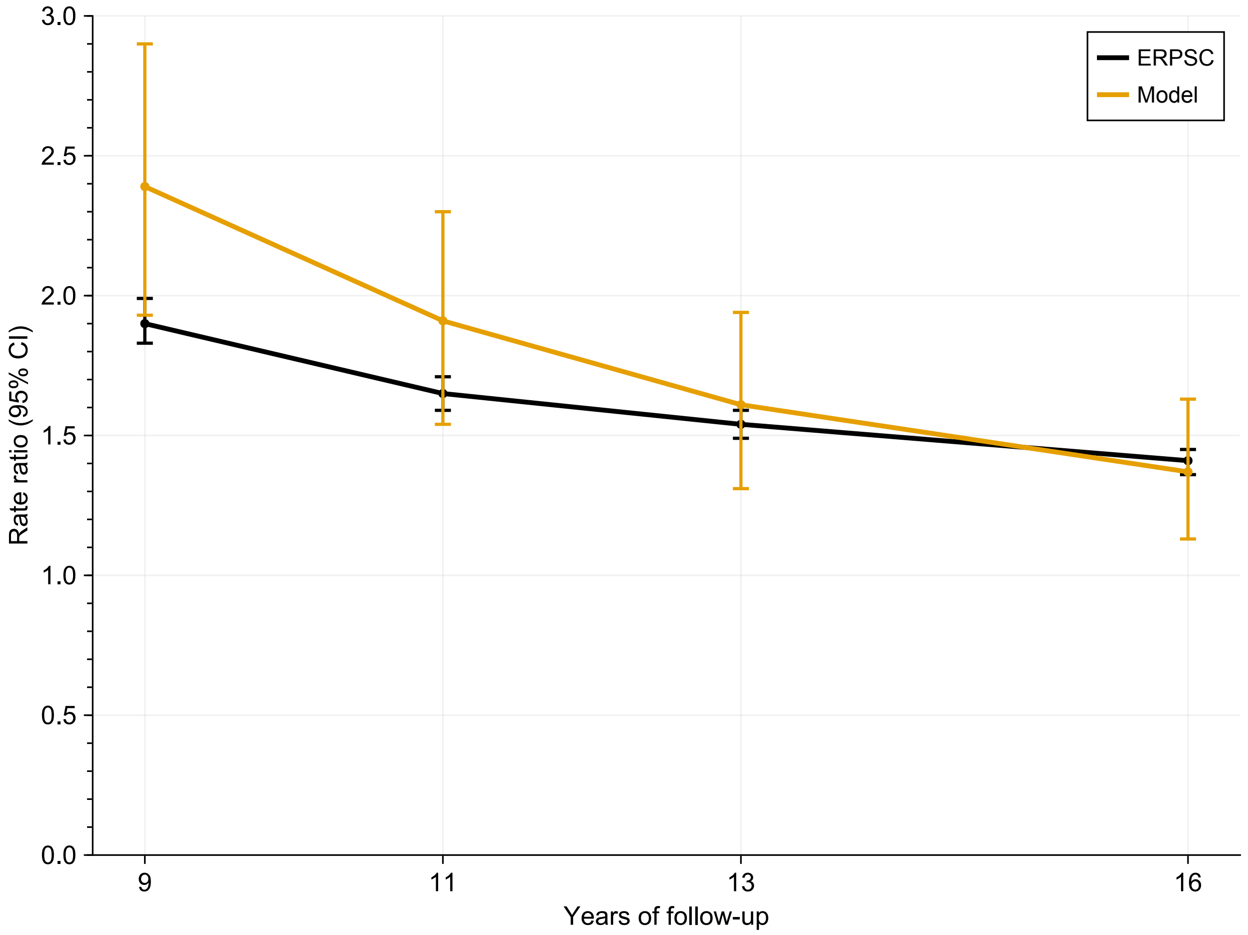


**Appendix Figure B.1:** Plot showing the rate ratio of prostate cancers detected over 9, 11, 13, and 16 years using our model against that seen in the European Randomized Study of Prostate Cancer Screening (ERPSC). Note, our model follows a single cohort from age 55 onwards, whereas the ERSPC involves a mixed cohort. Key parameters were MST of 12.6 (95% intervals: 11.2–14.2) years and episode sensitivity of 59%.

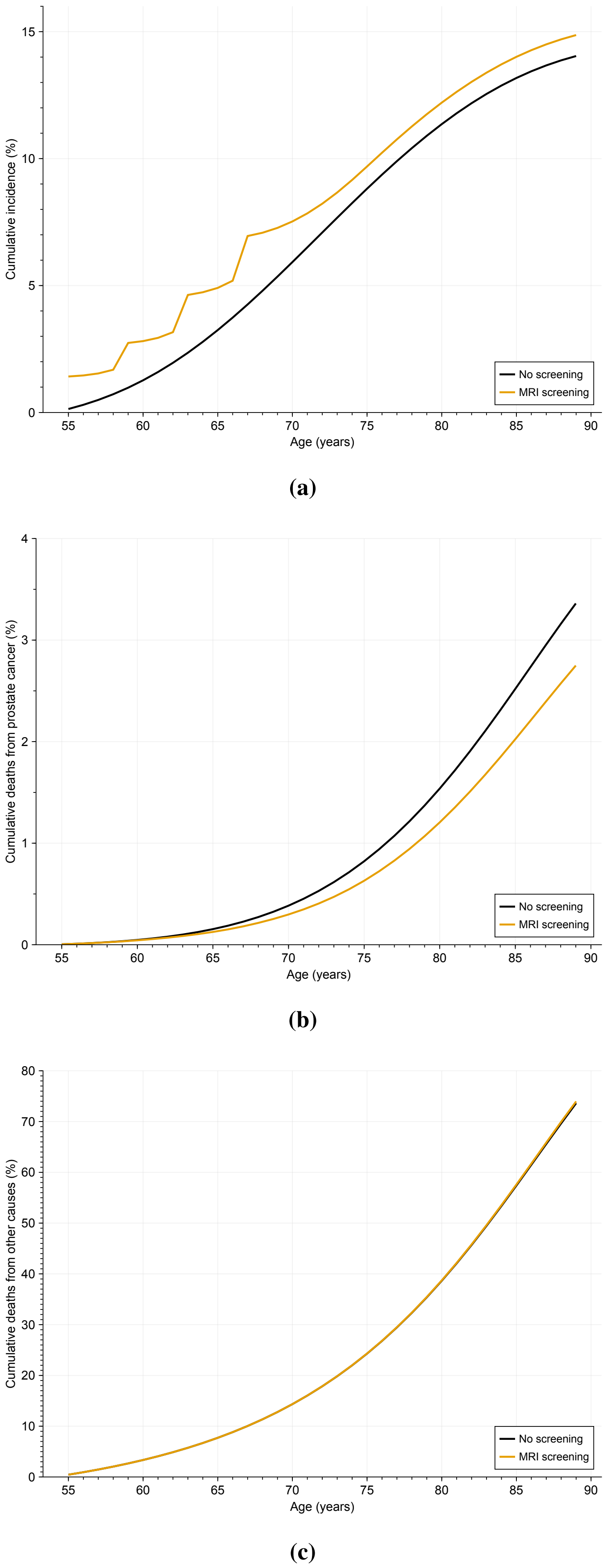


**Appendix Figure B.2:** Cumulative percentage with prostate cancer (A), dying from prostate cancer (B), and dying from other causes (C) by age for no screening and MRI screening cohorts.

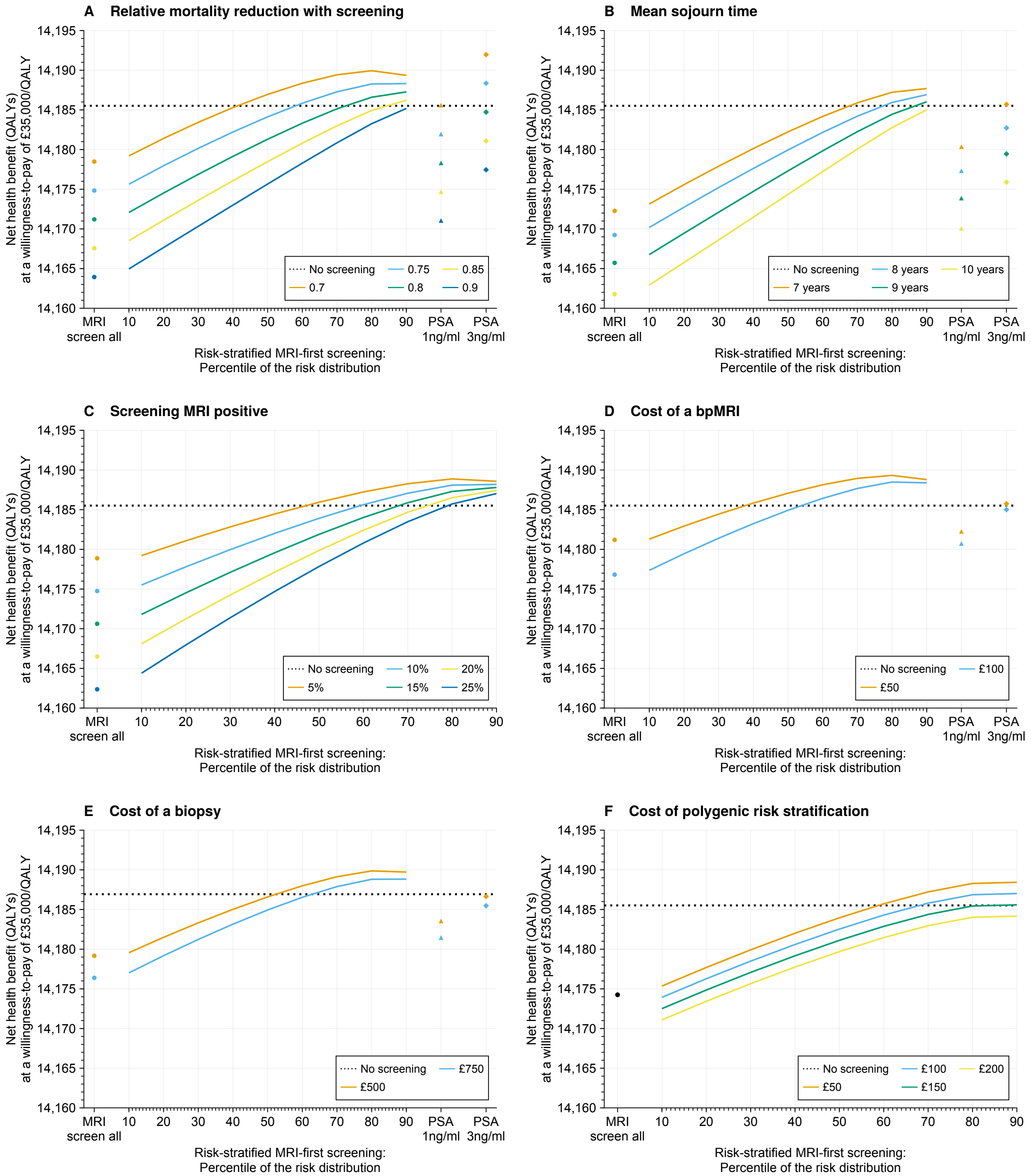


**Appendix Figure B.3:** Scenario analysis: Impact of varying different parameters on the net health benefit of age-based and risk-stratified MRI screening every four years from ages 55–69 at a willingness-to-pay of £35,000 per QALY gained. Abbreviations: bpMRI, biparametric MRI.

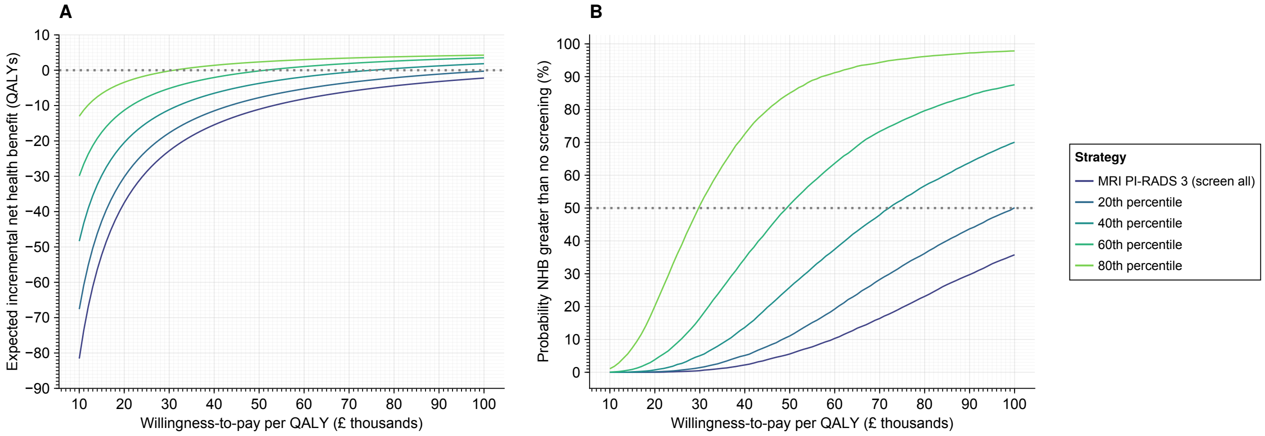


**Appendix Figure B.4:** Net health benefit (NHB) by willingness-to-pay (WTP) threshold for MRI-first screening where a PI-RADS score of $\geq 3$ is considered screen positive. (A) shows the expected incremental NHB of MRI-first screening relative to no screening, alongside (B) the probability of the incremental NHB being greater than that of no screening. The probability of having an incremental NHB > 0 provides a measure of the spread of outcomes relative to no screening. The strategy with the highest NHB at a given WTP would be considered most cost-effective.

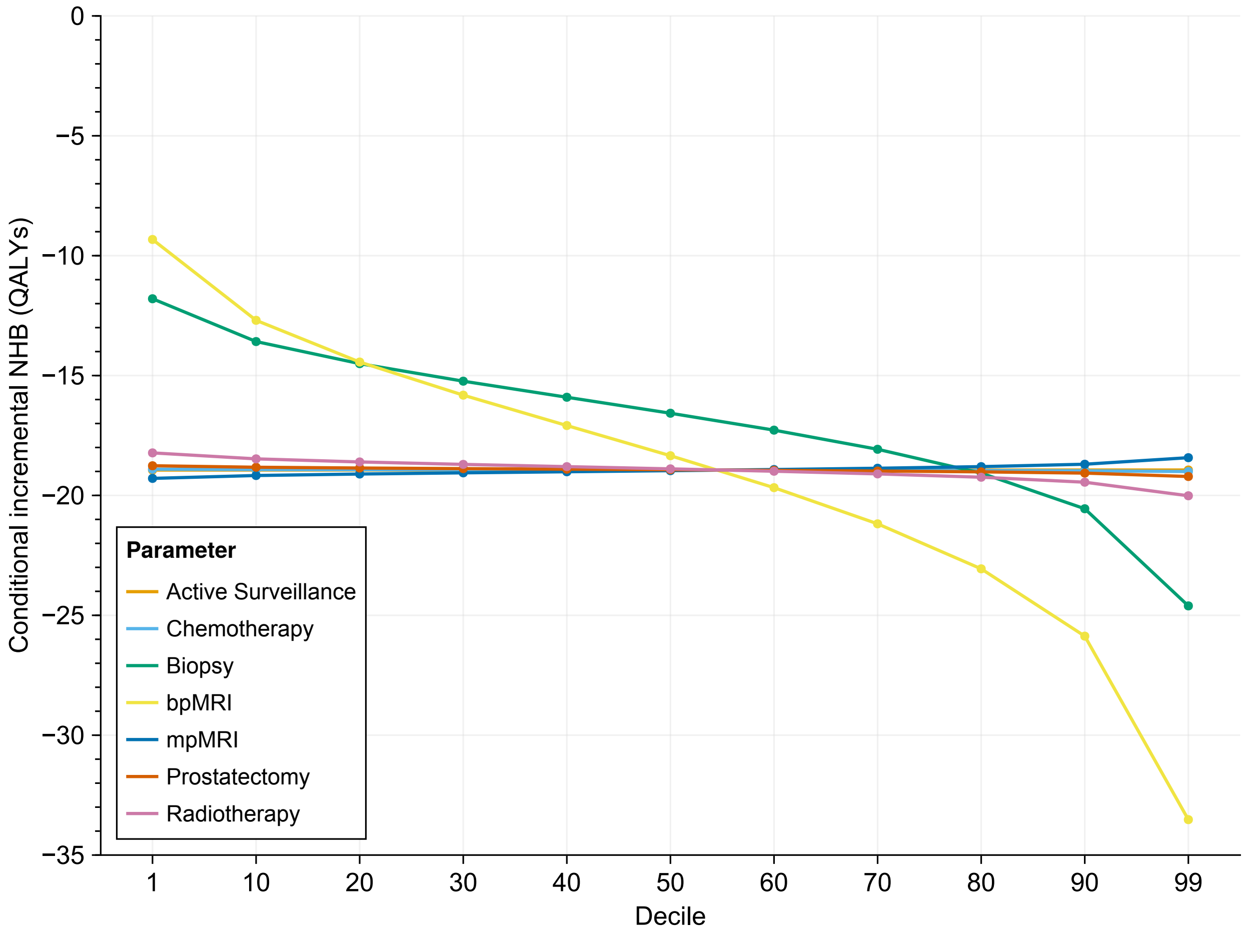


**Appendix Figure B.5:** Probabilistic one-way sensitivity analysis of MRI-first screening [1]. The cost-effectiveness of screening was most sensitive to the cost of biopsy and biparametric MRI. Net health benefit (NHB) calculated at a willingness-to-pay of £25,000 per quality-adjusted life-year (QALY). Underlying values in the table below. Abbreviations: bpMRI, biparametric MRI; mpMRI, multiparametric MRI.

Probabilistic one-way sensitivity analysis costs in GBP £ for MRI-first screening

| | Decile | | | | | | | | | | |
|---|---|---|---|---|---|---|---|---|---|---|---|
| Variable | 1 | 10 | 20 | 30 | 40 | 50 | 60 | 70 | 80 | 90 | 99 |
| Active surveillance | 2,509 | 3,865 | 4,565 | 5,121 | 5,630 | 6,136 | 6,672 | 7,279 | 8,034 | 9,163 | 12,240 |
| Chemotherapy | 1,019 | 1,570 | 1,854 | 2,080 | 2,287 | 2,493 | 2,710 | 2,957 | 3,263 | 3,722 | 4,972 |
| Biopsy | 298 | 458 | 541 | 607 | 668 | 728 | 791 | 863 | 953 | 1,087 | 1,452 |
| bpMRI | 51 | 78 | 93 | 104 | 114 | 124 | 135 | 147 | 163 | 186 | 248 |
| mpMRI | 78 | 120 | 142 | 159 | 175 | 191 | 208 | 226 | 250 | 285 | 381 |
| Prostatectomy | 4,438 | 6,836 | 8,073 | 9,056 | 9,957 | 10,852 | 11,800 | 12,872 | 14,207 | 16,204 | 21,647 |
| Radiotherapy | 3,055 | 4,705 | 5,557 | 6,233 | 6,853 | 7,470 | 8,122 | 8,860 | 9,779 | 11,153 | 14,900 |

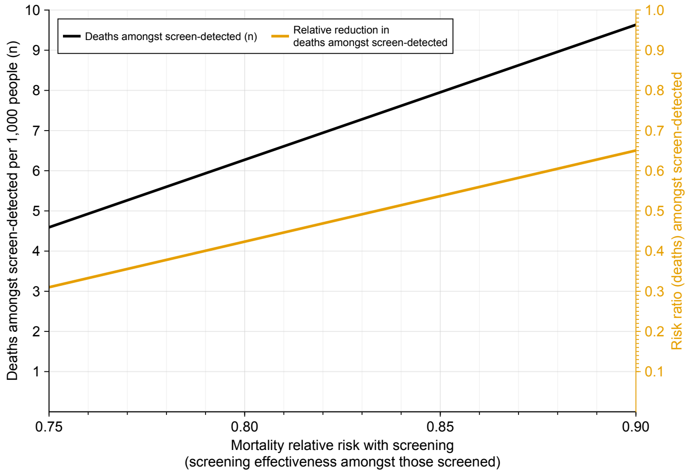


**(a)** MRI-first screening

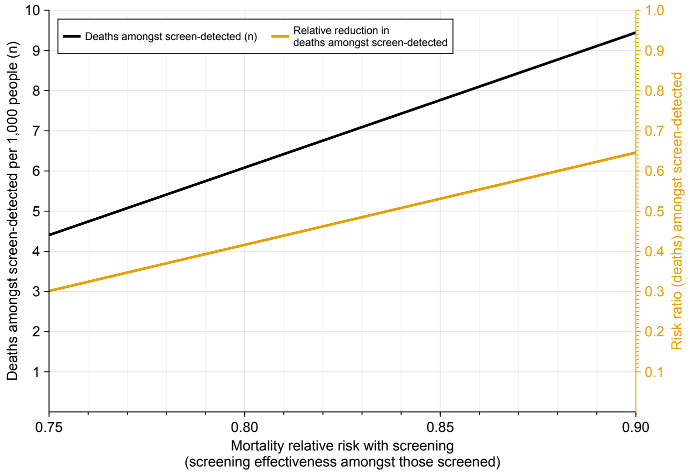


**(b)** PSA (1ng/ml threshold)

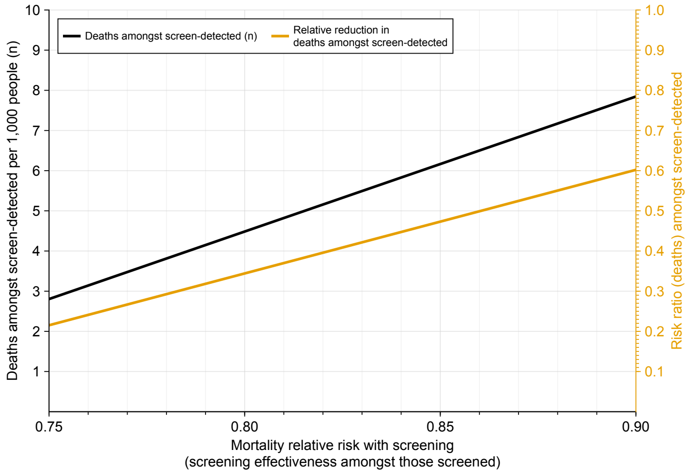


**(c)** PSA (3ng/ml threshold)

**Appendix Figure B.6:** Necessary mortality reductions amongst those whose cancer was screen-detected to observe a particular relative reduction amongst all those who received screening. For example, for MRI-first screening (subfigure A), if the screening efficacy is 20% (i.e. a relative risk of 0.8 in cancer deaths amongst all those screened), deaths amongst those whose cancer was screen-detected would need to be reduced by approximately 58%.

**Appendix Table B.1:** Outcomes for risk-stratified MRI-first screening strategies every 4 years from age 55–69 with follow-up to age 89 per 1,000 men

| Strategy | Prostate cancers | Screen-detected cancers | Interval cancers | Over-diagnosed cancers | Deaths from prostate cancer | Screening MRI | Diagnostic MRI | Total MRI | Biopsies | Life-years | QALYs | Costs (£) | WTP threshold* |
|---|---|---|---|---|---|---|---|---|---|---|---|---|---|
| No screening | 140 | 0 | 0 | 0 | 34 | 0 | 263 | 263 | 176 | 16,965 | 14,211 | 878,516 | N/A |
| MRI-first | 149 | 57 | 13 | 8 | 27 | 3,693 | 172 | 3,865 | 507 | 16,984 | 14,219 | 1,550,738 | 82,000 |
| *Polygenic risk centiles* | | | | | | | | | | | | | |
| 1 | 149 | 57 | 12 | 8 | 27 | 3,655 | 172 | 3,827 | 503 | 16,984 | 14,219 | 1,602,820 | 88,000 |
| 2 | 148 | 57 | 12 | 8 | 27 | 3,617 | 172 | 3,789 | 499 | 16,984 | 14,219 | 1,595,119 | 86,000 |
| 3 | 148 | 57 | 12 | 8 | 27 | 3,579 | 172 | 3,751 | 495 | 16,984 | 14,219 | 1,587,442 | 85,000 |
| 4 | 148 | 57 | 12 | 8 | 27 | 3,541 | 172 | 3,713 | 490 | 16,984 | 14,219 | 1,579,784 | 84,000 |
| 5 | 148 | 57 | 12 | 8 | 27 | 3,503 | 172 | 3,674 | 486 | 16,984 | 14,219 | 1,572,143 | 82,000 |
| 6 | 148 | 56 | 12 | 8 | 27 | 3,465 | 172 | 3,637 | 482 | 16,984 | 14,219 | 1,564,516 | 81,000 |
| 7 | 148 | 56 | 12 | 8 | 27 | 3,427 | 172 | 3,599 | 478 | 16,984 | 14,219 | 1,556,902 | 80,000 |
| 8 | 148 | 56 | 12 | 7 | 27 | 3,389 | 172 | 3,561 | 474 | 16,984 | 14,219 | 1,549,300 | 79,000 |
| 9 | 148 | 56 | 12 | 7 | 27 | 3,351 | 172 | 3,523 | 470 | 16,984 | 14,219 | 1,541,709 | 77,000 |
| 10 | 148 | 56 | 12 | 7 | 27 | 3,313 | 172 | 3,485 | 467 | 16,984 | 14,219 | 1,534,128 | 76,000 |
| 11 | 147 | 56 | 12 | 7 | 27 | 3,275 | 172 | 3,447 | 463 | 16,984 | 14,219 | 1,526,557 | 75,000 |
| 12 | 147 | 55 | 12 | 7 | 27 | 3,237 | 172 | 3,409 | 459 | 16,984 | 14,219 | 1,518,996 | 74,000 |
| 13 | 147 | 55 | 12 | 7 | 27 | 3,199 | 172 | 3,372 | 455 | 16,984 | 14,219 | 1,511,444 | 73,000 |
| 14 | 147 | 55 | 12 | 7 | 27 | 3,161 | 173 | 3,334 | 451 | 16,984 | 14,219 | 1,503,900 | 72,000 |
| 15 | 147 | 55 | 12 | 7 | 27 | 3,123 | 173 | 3,296 | 447 | 16,984 | 14,219 | 1,496,365 | 71,000 |
| 16 | 147 | 55 | 12 | 7 | 27 | 3,086 | 173 | 3,258 | 443 | 16,984 | 14,219 | 1,488,837 | 70,000 |
| 17 | 147 | 55 | 12 | 7 | 27 | 3,048 | 173 | 3,221 | 439 | 16,984 | 14,219 | 1,481,318 | 69,000 |
| 18 | 147 | 54 | 12 | 7 | 27 | 3,010 | 173 | 3,183 | 435 | 16,984 | 14,219 | 1,473,807 | 68,000 |
| 19 | 146 | 54 | 12 | 7 | 27 | 2,972 | 173 | 3,146 | 431 | 16,984 | 14,219 | 1,466,304 | 67,000 |
| 20 | 146 | 54 | 12 | 7 | 27 | 2,934 | 173 | 3,108 | 427 | 16,984 | 14,219 | 1,458,808 | 66,000 |
| 21 | 146 | 54 | 12 | 7 | 27 | 2,897 | 174 | 3,070 | 423 | 16,984 | 14,219 | 1,451,320 | 65,000 |
| 22 | 146 | 53 | 12 | 7 | 27 | 2,859 | 174 | 3,033 | 420 | 16,984 | 14,219 | 1,443,839 | 64,000 |
| 23 | 146 | 53 | 12 | 7 | 27 | 2,821 | 174 | 2,995 | 416 | 16,984 | 14,219 | 1,436,366 | 63,000 |
| 24 | 146 | 53 | 12 | 7 | 27 | 2,783 | 174 | 2,958 | 412 | 16,984 | 14,219 | 1,428,900 | 62,000 |
| 25 | 146 | 53 | 12 | 7 | 28 | 2,746 | 175 | 2,920 | 408 | 16,983 | 14,219 | 1,421,441 | 61,000 |
| 26 | 146 | 52 | 11 | 7 | 28 | 2,708 | 175 | 2,883 | 404 | 16,983 | 14,219 | 1,413,990 | 60,000 |
| 27 | 146 | 52 | 11 | 7 | 28 | 2,670 | 175 | 2,846 | 400 | 16,983 | 14,219 | 1,406,547 | 60,000 |
| 28 | 145 | 52 | 11 | 7 | 28 | 2,633 | 175 | 2,808 | 397 | 16,983 | 14,219 | 1,399,110 | 59,000 |
| 29 | 145 | 52 | 11 | 7 | 28 | 2,595 | 176 | 2,771 | 393 | 16,983 | 14,219 | 1,391,682 | 58,000 |
| 30 | 145 | 51 | 11 | 7 | 28 | 2,557 | 176 | 2,733 | 389 | 16,983 | 14,219 | 1,384,260 | 57,000 |
| 31 | 145 | 51 | 11 | 7 | 28 | 2,520 | 176 | 2,696 | 385 | 16,983 | 14,219 | 1,376,847 | 56,000 |
| 32 | 145 | 51 | 11 | 7 | 28 | 2,482 | 177 | 2,659 | 382 | 16,983 | 14,219 | 1,369,441 | 55,000 |
| 33 | 145 | 50 | 11 | 7 | 28 | 2,445 | 177 | 2,622 | 378 | 16,983 | 14,219 | 1,362,042 | 54,000 |
| 34 | 145 | 50 | 11 | 7 | 28 | 2,407 | 177 | 2,584 | 374 | 16,983 | 14,219 | 1,354,652 | 54,000 |
| 35 | 145 | 50 | 11 | 7 | 28 | 2,370 | 178 | 2,547 | 370 | 16,983 | 14,219 | 1,347,269 | 53,000 |

*Continued on the next page*

**Appendix Table B.1**: Outcomes for risk-stratified MRI-first screening strategies every 4 years from age 55–69 with follow-up to age 89 per 1,000 men, continued

| Strategy | Prostate cancers | Screen-detected cancers | Interval cancers | Over-diagnosed cancers | Deaths from prostate cancer | Screening MRI | Diagnostic MRI | Total MRI | Biopsies | Life-years | QALYs | Costs (£) | WTP threshold* |
|---|---|---|---|---|---|---|---|---|---|---|---|---|---|
| 36 | 144 | 49 | 11 | 7 | 28 | 2,332 | 178 | 2,510 | 367 | 16,983 | 14,219 | 1,339,894 | 52,000 |
| 37 | 144 | 49 | 11 | 7 | 28 | 2,294 | 178 | 2,473 | 363 | 16,983 | 14,219 | 1,332,528 | 51,000 |
| 38 | 144 | 49 | 11 | 7 | 28 | 2,257 | 179 | 2,436 | 359 | 16,983 | 14,219 | 1,325,170 | 50,000 |
| 39 | 144 | 48 | 11 | 6 | 28 | 2,219 | 179 | 2,399 | 355 | 16,983 | 14,219 | 1,317,820 | 50,000 |
| 40 | 144 | 48 | 11 | 6 | 28 | 2,182 | 180 | 2,362 | 352 | 16,983 | 14,219 | 1,310,478 | 49,000 |
| 41 | 144 | 48 | 10 | 6 | 28 | 2,145 | 180 | 2,325 | 348 | 16,982 | 14,219 | 1,303,146 | 48,000 |
| 42 | 144 | 47 | 10 | 6 | 28 | 2,107 | 180 | 2,288 | 344 | 16,982 | 14,219 | 1,295,822 | 47,000 |
| 43 | 143 | 47 | 10 | 6 | 28 | 2,070 | 181 | 2,251 | 341 | 16,982 | 14,219 | 1,288,508 | 46,000 |
| 44 | 143 | 47 | 10 | 6 | 28 | 2,032 | 181 | 2,214 | 337 | 16,982 | 14,219 | 1,281,203 | 46,000 |
| 45 | 143 | 46 | 10 | 6 | 28 | 1,995 | 182 | 2,177 | 333 | 16,982 | 14,219 | 1,273,907 | 45,000 |
| 46 | 143 | 46 | 10 | 6 | 28 | 1,958 | 182 | 2,140 | 330 | 16,982 | 14,219 | 1,266,621 | 44,000 |
| 47 | 143 | 45 | 10 | 6 | 28 | 1,920 | 183 | 2,103 | 326 | 16,982 | 14,219 | 1,259,345 | 43,000 |
| 48 | 143 | 45 | 10 | 6 | 28 | 1,883 | 183 | 2,066 | 322 | 16,982 | 14,219 | 1,252,080 | 43,000 |
| 49 | 143 | 45 | 10 | 6 | 28 | 1,846 | 184 | 2,030 | 319 | 16,982 | 14,219 | 1,244,825 | 42,000 |
| 50 | 143 | 44 | 10 | 6 | 28 | 1,808 | 184 | 1,993 | 315 | 16,982 | 14,219 | 1,237,582 | 41,000 |
| 51 | 142 | 44 | 10 | 6 | 28 | 1,771 | 185 | 1,956 | 312 | 16,981 | 14,219 | 1,230,349 | 41,000 |
| 52 | 142 | 43 | 9 | 6 | 28 | 1,734 | 186 | 1,919 | 308 | 16,981 | 14,219 | 1,223,128 | 40,000 |
| 53 | 142 | 43 | 9 | 6 | 28 | 1,697 | 186 | 1,883 | 305 | 16,981 | 14,219 | 1,215,920 | 39,000 |
| 54 | 142 | 42 | 9 | 6 | 28 | 1,659 | 187 | 1,846 | 301 | 16,981 | 14,219 | 1,208,723 | 38,000 |
| 55 | 142 | 42 | 9 | 6 | 28 | 1,622 | 187 | 1,810 | 298 | 16,981 | 14,219 | 1,201,540 | 38,000 |
| 56 | 142 | 41 | 9 | 6 | 28 | 1,585 | 188 | 1,773 | 294 | 16,981 | 14,219 | 1,194,370 | 37,000 |
| 57 | 142 | 41 | 9 | 5 | 28 | 1,548 | 189 | 1,737 | 291 | 16,981 | 14,219 | 1,187,214 | 36,000 |
| 58 | 141 | 40 | 9 | 5 | 28 | 1,511 | 189 | 1,700 | 287 | 16,981 | 14,219 | 1,180,073 | 36,000 |
| 59 | 141 | 40 | 9 | 5 | 28 | 1,474 | 190 | 1,664 | 284 | 16,980 | 14,219 | 1,172,946 | 35,000 |
| 60 | 141 | 39 | 9 | 5 | 28 | 1,437 | 191 | 1,628 | 280 | 16,980 | 14,219 | 1,165,836 | 34,000 |
| 61 | 141 | 39 | 8 | 5 | 28 | 1,400 | 192 | 1,591 | 277 | 16,980 | 14,219 | 1,158,741 | 34,000 |
| 62 | 141 | 38 | 8 | 5 | 28 | 1,363 | 192 | 1,555 | 273 | 16,980 | 14,219 | 1,151,664 | 33,000 |
| 63 | 141 | 38 | 8 | 5 | 29 | 1,326 | 193 | 1,519 | 270 | 16,980 | 14,219 | 1,144,605 | 32,000 |
| 64 | 141 | 37 | 8 | 5 | 29 | 1,289 | 194 | 1,483 | 267 | 16,980 | 14,219 | 1,137,564 | 32,000 |
| 65 | 140 | 37 | 8 | 5 | 29 | 1,252 | 195 | 1,447 | 263 | 16,980 | 14,219 | 1,130,544 | 31,000 |
| 66 | 140 | 36 | 8 | 5 | 29 | 1,215 | 196 | 1,411 | 260 | 16,979 | 14,218 | 1,123,543 | 30,000 |
| 67 | 140 | 35 | 8 | 5 | 29 | 1,178 | 196 | 1,375 | 257 | 16,979 | 14,218 | 1,116,565 | 30,000 |
| 68 | 140 | 35 | 8 | 5 | 29 | 1,141 | 197 | 1,339 | 253 | 16,979 | 14,218 | 1,109,610 | 29,000 |
| 69 | 140 | 34 | 7 | 5 | 29 | 1,104 | 198 | 1,303 | 250 | 16,979 | 14,218 | 1,102,679 | 28,000 |
| 70 | 140 | 34 | 7 | 4 | 29 | 1,068 | 199 | 1,267 | 247 | 16,979 | 14,218 | 1,095,774 | 28,000 |
| 71 | 140 | 33 | 7 | 4 | 29 | 1,031 | 200 | 1,231 | 243 | 16,978 | 14,218 | 1,088,896 | 27,000 |
| 72 | 139 | 32 | 7 | 4 | 29 | 994 | 201 | 1,196 | 240 | 16,978 | 14,218 | 1,082,047 | 26,000 |
| 73 | 139 | 31 | 7 | 4 | 29 | 958 | 202 | 1,160 | 237 | 16,978 | 14,218 | 1,075,228 | 26,000 |
| 74 | 139 | 31 | 7 | 4 | 29 | 921 | 203 | 1,124 | 234 | 16,978 | 14,218 | 1,068,443 | 25,000 |

*Continued on the next page*

**Appendix Table B.1**: Outcomes for risk-stratified MRI-first screening strategies every 4 years from age 55–69 with follow-up to age 89 per 1,000 men, continued

| Strategy | Prostate cancers | Screen-detected cancers | Interval cancers | Over-diagnosed cancers | Deaths from prostate cancer | Screening MRI | Diagnostic MRI | Total MRI | Biopsies | Life-years | QALYs | Costs (£) | WTP threshold* |
|---|---|---|---|---|---|---|---|---|---|---|---|---|---|
| 75 | 139 | 30 | 7 | 4 | 29 | 885 | 204 | 1,089 | 231 | 16,978 | 14,218 | 1,061,693 | 25,000 |
| 76 | 139 | 29 | 6 | 4 | 29 | 848 | 206 | 1,054 | 228 | 16,977 | 14,218 | 1,054,981 | 24,000 |
| 77 | 139 | 29 | 6 | 4 | 29 | 812 | 207 | 1,018 | 225 | 16,977 | 14,218 | 1,048,309 | 23,000 |
| 78 | 139 | 28 | 6 | 4 | 29 | 775 | 208 | 983 | 222 | 16,977 | 14,218 | 1,041,682 | 23,000 |
| 79 | 139 | 27 | 6 | 4 | 30 | 739 | 209 | 948 | 218 | 16,977 | 14,217 | 1,035,101 | 22,000 |
| 80 | 138 | 26 | 6 | 3 | 30 | 702 | 211 | 913 | 216 | 16,976 | 14,217 | 1,028,573 | 22,000 |
| 81 | 138 | 25 | 5 | 3 | 30 | 666 | 212 | 878 | 213 | 16,976 | 14,217 | 1,022,100 | 21,000 |
| 82 | 138 | 24 | 5 | 3 | 30 | 630 | 213 | 843 | 210 | 16,976 | 14,217 | 1,015,689 | 21,000 |
| 83 | 138 | 24 | 5 | 3 | 30 | 594 | 215 | 809 | 207 | 16,975 | 14,217 | 1,009,345 | 20,000 |
| 84 | 138 | 23 | 5 | 3 | 30 | 558 | 216 | 774 | 204 | 16,975 | 14,217 | 1,003,076 | 20,000 |
| 85 | 138 | 22 | 5 | 3 | 30 | 522 | 218 | 740 | 201 | 16,975 | 14,217 | 996,890 | 19,000 |
| 86 | 138 | 21 | 4 | 3 | 30 | 486 | 220 | 705 | 199 | 16,974 | 14,217 | 990,796 | 19,000 |
| 87 | 138 | 20 | 4 | 3 | 30 | 450 | 221 | 671 | 196 | 16,974 | 14,216 | 984,807 | 18,000 |
| 88 | 138 | 19 | 4 | 2 | 31 | 414 | 223 | 637 | 193 | 16,974 | 14,216 | 978,935 | 18,000 |
| 89 | 137 | 17 | 4 | 2 | 31 | 379 | 225 | 604 | 191 | 16,973 | 14,216 | 973,199 | 17,000 |
| 90 | 137 | 16 | 4 | 2 | 31 | 343 | 227 | 570 | 189 | 16,973 | 14,216 | 967,618 | 17,000 |
| 91 | 137 | 15 | 3 | 2 | 31 | 308 | 229 | 537 | 186 | 16,972 | 14,216 | 962,219 | 17,000 |
| 92 | 137 | 14 | 3 | 2 | 31 | 272 | 232 | 504 | 184 | 16,972 | 14,215 | 957,036 | 17,000 |
| 93 | 137 | 13 | 3 | 2 | 31 | 237 | 234 | 471 | 182 | 16,971 | 14,215 | 952,113 | 17,000 |
| 94 | 137 | 11 | 2 | 1 | 31 | 202 | 237 | 439 | 180 | 16,971 | 14,215 | 947,510 | 17,000 |
| 95 | 138 | 10 | 2 | 1 | 32 | 168 | 240 | 407 | 178 | 16,970 | 14,214 | 943,312 | 17,000 |
| 96 | 138 | 8 | 2 | 1 | 32 | 133 | 243 | 376 | 177 | 16,969 | 14,214 | 939,644 | 19,000 |
| 97 | 138 | 6 | 1 | 1 | 32 | 99 | 247 | 346 | 176 | 16,969 | 14,213 | 936,705 | 21,000 |
| 98 | 138 | 5 | 1 | 1 | 33 | 65 | 251 | 316 | 175 | 16,968 | 14,213 | 934,841 | 27,000 |
| 99 | 139 | 2 | 1 | 0 | 33 | 32 | 256 | 288 | 175 | 16,967 | 14,212 | 934,768 | 44,000 |

Main analyses. All strategies followed 1,000 individuals from age 55 through 89. Where relevant, screening was every 4 years between 55 and 69. Each centile of the risk distribution is shown.

* Willingness-to-pay (WTP) threshold needed for the strategy to have a >50% probability of having an incremental net health benefit (NHB) greater than that of no screening. For context, the UK National Institute for Health and Care Excellence set thresholds for most interventions at between £25,000–£35,000 to be considered cost-effective.

Abbreviations: MRI, magnetic resonance imaging; QALYs, quality-adjusted life-years; PSA, prostate specific antigen.

**Appendix Table B.2:** Overdiagnosis estimated using excess incidence and lead time methods

| Strategy | Overdiagnosis: excess incidence (%)[a] | Overdiagnosis: lead time (%) |
|---|---|---|
| MRI-first | 14.2 (2.3–25.3) | 13.3 (10.8–16.0) |
| PSA, 1ng/ml | 16.9 (7.2–26.7) | 15.2 (12.2–18.4) |
| PSA, 3ng/ml | 18.2 (9.2–27.4) | 15.4 (12.3–18.7) |

[a] Excess incidence calculated as the difference in cumulative incidence at age 89 by strategy compared with no screening.

All strategies involve screening every four years from aged 55–69, with the cohort followed to age 89. Percentages refer to overdiagnosis relative to the number of screen-detected cancers. Values are means with 95% intervals across the probabilistic runs.

**Appendix Table B.3:** Screening strategies ranked by the probability that their incremental net health benefit exceeds zero at £25,000 per QALY

| Strategy | QALYs | Costs (£) | NHB (£25k) | iNHB > 0 (%) | Rank |
|---|---|---|---|---|---|
| No screening | 14,211 | 878,516 | 14,175 | | |
| MRI-first | 14,219 | 1,550,738 | 14,157 | 1.7 | 95 |
| *Polygenic risk centiles* | | | | | |
| 1 | 14,219 | 1,602,820 | 14,154 | 1.2 | 102 |
| 2 | 14,219 | 1,595,119 | 14,155 | 1.2 | 101 |
| 3 | 14,219 | 1,587,442 | 14,155 | 1.3 | 100 |
| 4 | 14,219 | 1,579,784 | 14,156 | 1.4 | 99 |
| 5 | 14,219 | 1,572,143 | 14,156 | 1.5 | 98 |
| 6 | 14,219 | 1,564,516 | 14,156 | 1.6 | 97 |
| 7 | 14,219 | 1,556,902 | 14,157 | 1.7 | 96 |
| 8 | 14,219 | 1,549,300 | 14,157 | 1.8 | 94 |
| 9 | 14,219 | 1,541,709 | 14,157 | 1.9 | 93 |
| 10 | 14,219 | 1,534,128 | 14,158 | 2.1 | 92 |
| 11 | 14,219 | 1,526,557 | 14,158 | 2.2 | 91 |
| 12 | 14,219 | 1,518,996 | 14,158 | 2.3 | 90 |
| 13 | 14,219 | 1,511,444 | 14,159 | 2.5 | 89 |
| 14 | 14,219 | 1,503,900 | 14,159 | 2.6 | 88 |
| 15 | 14,219 | 1,496,365 | 14,159 | 2.7 | 87 |
| 16 | 14,219 | 1,488,837 | 14,159 | 2.9 | 86 |
| 17 | 14,219 | 1,481,318 | 14,160 | 3.0 | 85 |
| 18 | 14,219 | 1,473,807 | 14,160 | 3.2 | 84 |
| 19 | 14,219 | 1,466,304 | 14,160 | 3.3 | 83 |
| 20 | 14,219 | 1,458,808 | 14,161 | 3.4 | 82 |
| 21 | 14,219 | 1,451,320 | 14,161 | 3.7 | 81 |
| 22 | 14,219 | 1,443,839 | 14,161 | 3.9 | 80 |
| 23 | 14,219 | 1,436,366 | 14,162 | 4.1 | 79 |
| 24 | 14,219 | 1,428,900 | 14,162 | 4.4 | 78 |
| 25 | 14,219 | 1,421,441 | 14,162 | 4.6 | 77 |
| 26 | 14,219 | 1,413,990 | 14,163 | 4.9 | 76 |
| 27 | 14,219 | 1,406,547 | 14,163 | 5.2 | 75 |
| 28 | 14,219 | 1,399,110 | 14,163 | 5.4 | 74 |
| 29 | 14,219 | 1,391,682 | 14,164 | 5.6 | 73 |
| 30 | 14,219 | 1,384,260 | 14,164 | 5.9 | 72 |
| 31 | 14,219 | 1,376,847 | 14,164 | 6.3 | 71 |
| 32 | 14,219 | 1,369,441 | 14,164 | 6.8 | 70 |
| 33 | 14,219 | 1,362,042 | 14,165 | 7.0 | 68 |
| 34 | 14,219 | 1,354,652 | 14,165 | 7.4 | 67 |
| 35 | 14,219 | 1,347,269 | 14,165 | 7.8 | 65 |
| 36 | 14,219 | 1,339,894 | 14,166 | 8.4 | 64 |
| 37 | 14,219 | 1,332,528 | 14,166 | 8.9 | 63 |
| 38 | 14,219 | 1,325,170 | 14,166 | 9.4 | 62 |
| 39 | 14,219 | 1,317,820 | 14,166 | 9.8 | 61 |
| 40 | 14,219 | 1,310,478 | 14,167 | 10.2 | 60 |
| 41 | 14,219 | 1,303,146 | 14,167 | 10.7 | 59 |
| 42 | 14,219 | 1,295,822 | 14,167 | 11.2 | 58 |
| 43 | 14,219 | 1,288,508 | 14,168 | 11.8 | 57 |
| 44 | 14,219 | 1,281,203 | 14,168 | 12.3 | 56 |
| 45 | 14,219 | 1,273,907 | 14,168 | 13.0 | 55 |
| 46 | 14,219 | 1,266,621 | 14,168 | 13.6 | 54 |
| 47 | 14,219 | 1,259,345 | 14,169 | 14.3 | 53 |
| 48 | 14,219 | 1,252,080 | 14,169 | 15.1 | 52 |
| 49 | 14,219 | 1,244,825 | 14,169 | 15.8 | 51 |
| 50 | 14,219 | 1,237,582 | 14,170 | 16.6 | 50 |
| 51 | 14,219 | 1,230,349 | 14,170 | 17.5 | 49 |
| 52 | 14,219 | 1,223,128 | 14,170 | 18.6 | 48 |
| 53 | 14,219 | 1,215,920 | 14,170 | 19.6 | 47 |
| 54 | 14,219 | 1,208,723 | 14,171 | 20.5 | 46 |
| 55 | 14,219 | 1,201,540 | 14,171 | 21.6 | 45 |
| 56 | 14,219 | 1,194,370 | 14,171 | 22.7 | 44 |
| 57 | 14,219 | 1,187,214 | 14,171 | 23.8 | 43 |
| 58 | 14,219 | 1,180,073 | 14,172 | 25.0 | 42 |

*Continued on the next page*

**Appendix Table B.3**: Screening strategies ranked by the probability that their incremental net health benefit relative to no screening exceeds zero at £25,000 per QALY

| Strategy | QALYs | Costs (£) | NHB (£25k) | iNHB > 0 (%) | Rank |
|---|---|---|---|---|---|
| 59 | 14,219 | 1,172,946 | 14,172 | 26.2 | 41 |
| 60 | 14,219 | 1,165,836 | 14,172 | 27.3 | 39 |
| 61 | 14,219 | 1,158,741 | 14,172 | 28.5 | 38 |
| 62 | 14,219 | 1,151,664 | 14,173 | 29.6 | 37 |
| 63 | 14,219 | 1,144,605 | 14,173 | 31.0 | 36 |
| 64 | 14,219 | 1,137,564 | 14,173 | 32.5 | 35 |
| 65 | 14,219 | 1,130,544 | 14,173 | 34.1 | 34 |
| 66 | 14,218 | 1,123,543 | 14,174 | 35.6 | 33 |
| 67 | 14,218 | 1,116,565 | 14,174 | 37.3 | 32 |
| 68 | 14,218 | 1,109,610 | 14,174 | 39.2 | 31 |
| 69 | 14,218 | 1,102,679 | 14,174 | 41.1 | 30 |
| 70 | 14,218 | 1,095,774 | 14,174 | 42.9 | 29 |
| 71 | 14,218 | 1,088,896 | 14,175 | 45.0 | 28 |
| 72 | 14,218 | 1,082,047 | 14,175 | 46.8 | 26 |
| 73 | 14,218 | 1,075,228 | 14,175 | 48.6 | 25 |
| 74 | 14,218 | 1,068,443 | 14,175 | 50.8 | 24 |
| 75 | 14,218 | 1,061,693 | 14,175 | 52.9 | 23 |
| 76 | 14,218 | 1,054,981 | 14,176 | 54.9 | 22 |
| 77 | 14,218 | 1,048,309 | 14,176 | 56.9 | 21 |
| 78 | 14,218 | 1,041,682 | 14,176 | 59.4 | 20 |
| 79 | 14,217 | 1,035,101 | 14,176 | 61.4 | 19 |
| 80 | 14,217 | 1,028,573 | 14,176 | 63.6 | 18 |
| 81 | 14,217 | 1,022,100 | 14,176 | 65.8 | 17 |
| 82 | 14,217 | 1,015,689 | 14,177 | 68.0 | 16 |
| 83 | 14,217 | 1,009,345 | 14,177 | 70.0 | 14 |
| 84 | 14,217 | 1,003,076 | 14,177 | 72.3 | 13 |
| 85 | 14,217 | 996,890 | 14,177 | 74.1 | 12 |
| 86 | 14,217 | 990,796 | 14,177 | 75.8 | 11 |
| 87 | 14,216 | 984,807 | 14,177 | 77.8 | 10 |
| 88 | 14,216 | 978,935 | 14,177 | 79.5 | 8 |
| 89 | 14,216 | 973,199 | 14,177 | 81.0 | 7 |
| 90 | 14,216 | 967,618 | 14,177 | 82.3 | 6 |
| 91 | 14,216 | 962,219 | 14,177 | 83.5 | 4 |
| 92 | 14,215 | 957,036 | 14,177 | 84.4 | 3 |
| 93 | 14,215 | 952,113 | 14,177 | 84.8 | 1 |
| 94 | 14,215 | 947,510 | 14,177 | 84.5 | 2 |
| 95 | 14,214 | 943,312 | 14,177 | 83.1 | 5 |
| 96 | 14,214 | 939,644 | 14,176 | 79.1 | 9 |
| 97 | 14,213 | 936,705 | 14,176 | 69.9 | 15 |
| 98 | 14,213 | 934,841 | 14,175 | 45.2 | 27 |
| 99 | 14,212 | 934,768 | 14,174 | 7.8 | 65 |
| PSA, 1ng/ml | 14,216 | 1,269,439 | 14,165 | 6.9 | 69 |
| PSA, 3ng/ml | 14,215 | 1,074,011 | 14,172 | 26.9 | 40 |

Incremental NHB relative to no screening.

**Appendix Table B.4:** Scenario analysis: The impact of varying the relative impact of screening every 4 years from age 55–69 on prostate cancer-specific mortality

| Strategy | Prostate cancers | Screen-detected cancers | Interval cancers | Over-diagnosed cancers | Deaths from prostate cancer | Screening MRI | Diagnostic MRI | Total MRI | Biopsies | Life-years | QALYs | Costs (£) | WTP threshold* |
|---|---|---|---|---|---|---|---|---|---|---|---|---|---|
| No screening | 140 | 0 | 0 | 0 | 34 | 0 | 263 | 263 | 176 | 16,965 | 14,211 | 878,516 | N/A |
| **RR=0.7** | | | | | | | | | | | | | |
| MRI-first | 149 | 57 | 13 | 8 | 25 | 3,693 | 172 | 3,865 | 507 | 16,989 | 14,222 | 1,538,789 | 54,000 |
| *Polygenic risk-stratified screening* | | | | | | | | | | | | | |
| 10th centile | 148 | 56 | 12 | 7 | 25 | 3,313 | 172 | 3,485 | 467 | 16,989 | 14,223 | 1,522,433 | 52,000 |
| 20th centile | 146 | 54 | 12 | 7 | 25 | 2,934 | 173 | 3,108 | 427 | 16,989 | 14,223 | 1,447,516 | 46,000 |
| 30th centile | 145 | 51 | 11 | 7 | 25 | 2,557 | 176 | 2,733 | 389 | 16,988 | 14,223 | 1,373,502 | 40,000 |
| 40th centile | 144 | 48 | 11 | 6 | 26 | 2,182 | 180 | 2,362 | 352 | 16,987 | 14,222 | 1,300,389 | 35,000 |
| 50th centile | 143 | 44 | 10 | 6 | 26 | 1,808 | 184 | 1,993 | 315 | 16,986 | 14,222 | 1,228,314 | 30,000 |
| 60th centile | 141 | 39 | 9 | 5 | 27 | 1,437 | 191 | 1,628 | 280 | 16,984 | 14,221 | 1,157,573 | 25,000 |
| 70th centile | 140 | 34 | 7 | 4 | 28 | 1,068 | 199 | 1,267 | 247 | 16,982 | 14,221 | 1,088,746 | 21,000 |
| 80th centile | 138 | 26 | 6 | 3 | 29 | 702 | 211 | 913 | 216 | 16,979 | 14,219 | 1,023,097 | 17,000 |
| 90th centile | 137 | 16 | 4 | 2 | 30 | 343 | 227 | 570 | 189 | 16,974 | 14,217 | 964,204 | 14,000 |
| PSA, 1ng/ml | 150 | 58 | 14 | 9 | 25 | 1,290 | 173 | 1,463 | 392 | 16,989 | 14,221 | 1,255,147 | 34,000 |
| PSA, 3ng/ml | 150 | 52 | 16 | 8 | 25 | 621 | 184 | 805 | 235 | 16,989 | 14,222 | 1,055,798 | 15,000 |
| **RR=0.75** | | | | | | | | | | | | | |
| MRI-first | 149 | 57 | 13 | 8 | 27 | 3,693 | 172 | 3,865 | 507 | 16,985 | 14,219 | 1,547,888 | 76,000 |
| *Polygenic risk-stratified screening* | | | | | | | | | | | | | |
| 10th centile | 148 | 56 | 12 | 7 | 27 | 3,313 | 172 | 3,485 | 467 | 16,985 | 14,219 | 1,531,339 | 72,000 |
| 20th centile | 146 | 54 | 12 | 7 | 27 | 2,934 | 173 | 3,108 | 427 | 16,984 | 14,220 | 1,456,115 | 62,000 |
| 30th centile | 145 | 51 | 11 | 7 | 27 | 2,557 | 176 | 2,733 | 389 | 16,984 | 14,220 | 1,381,695 | 54,000 |
| 40th centile | 144 | 48 | 11 | 6 | 27 | 2,182 | 180 | 2,362 | 352 | 16,983 | 14,220 | 1,308,073 | 46,000 |
| 50th centile | 143 | 44 | 10 | 6 | 28 | 1,808 | 184 | 1,993 | 315 | 16,982 | 14,219 | 1,235,372 | 39,000 |
| 60th centile | 141 | 39 | 9 | 5 | 28 | 1,437 | 191 | 1,628 | 280 | 16,981 | 14,219 | 1,163,866 | 33,000 |
| 70th centile | 140 | 34 | 7 | 4 | 29 | 1,068 | 199 | 1,267 | 247 | 16,979 | 14,219 | 1,094,099 | 27,000 |
| 80th centile | 138 | 26 | 6 | 3 | 29 | 702 | 211 | 913 | 216 | 16,977 | 14,218 | 1,027,268 | 21,000 |
| 90th centile | 137 | 16 | 4 | 2 | 31 | 343 | 227 | 570 | 189 | 16,973 | 14,216 | 966,805 | 16,000 |
| PSA, 1ng/ml | 150 | 58 | 14 | 9 | 27 | 1,290 | 173 | 1,463 | 392 | 16,985 | 14,218 | 1,264,245 | 50,000 |

*Continued on the next page*

**Appendix Table B.4**: Scenario analysis: The impact of varying the relative impact of screening every 4 years from age 55–69 on prostate cancer-specific mortality, continued

| Strategy | Prostate cancers | Screen-detected cancers | Interval cancers | Over-diagnosed cancers | Deaths from prostate cancer | Screening MRI | Diagnostic MRI | Total MRI | Biopsies | Life-years | QALYs | Costs (£) | WTP threshold* |
|---|---|---|---|---|---|---|---|---|---|---|---|---|---|
| PSA, 3ng/ml | 150 | 52 | 16 | 8 | 27 | 621 | 184 | 805 | 235 | 16,985 | 14,219 | 1,064,597 | 22,000 |
| **RR=0.8** | | | | | | | | | | | | | |
| MRI-first | 149 | 57 | 13 | 8 | 29 | 3,693 | 172 | 3,865 | 507 | 16,980 | 14,216 | 1,556,967 | 127,000 |
| *Polygenic risk-stratified screening* | | | | | | | | | | | | | |
| 10th centile | 148 | 56 | 12 | 7 | 28 | 3,313 | 172 | 3,485 | 467 | 16,980 | 14,216 | 1,540,226 | 115,000 |
| 20th centile | 146 | 54 | 12 | 7 | 28 | 2,934 | 173 | 3,108 | 427 | 16,980 | 14,216 | 1,464,696 | 98,000 |
| 30th centile | 145 | 51 | 11 | 7 | 29 | 2,557 | 176 | 2,733 | 389 | 16,980 | 14,217 | 1,389,871 | 82,000 |
| 40th centile | 144 | 48 | 11 | 6 | 29 | 2,182 | 180 | 2,362 | 352 | 16,979 | 14,217 | 1,315,740 | 69,000 |
| 50th centile | 143 | 44 | 10 | 6 | 29 | 1,808 | 184 | 1,993 | 315 | 16,979 | 14,217 | 1,242,415 | 57,000 |
| 60th centile | 141 | 39 | 9 | 5 | 29 | 1,437 | 191 | 1,628 | 280 | 16,978 | 14,217 | 1,170,146 | 46,000 |
| 70th centile | 140 | 34 | 7 | 4 | 30 | 1,068 | 199 | 1,267 | 247 | 16,976 | 14,217 | 1,099,440 | 36,000 |
| 80th centile | 138 | 26 | 6 | 3 | 30 | 702 | 211 | 913 | 216 | 16,975 | 14,216 | 1,031,430 | 27,000 |
| 90th centile | 137 | 16 | 4 | 2 | 31 | 343 | 227 | 570 | 189 | 16,972 | 14,215 | 969,400 | 21,000 |
| PSA, 1ng/ml | 150 | 58 | 14 | 9 | 29 | 1,290 | 173 | 1,463 | 392 | 16,980 | 14,215 | 1,273,324 | 93,000 |
| PSA, 3ng/ml | 150 | 52 | 16 | 8 | 29 | 621 | 184 | 805 | 235 | 16,980 | 14,215 | 1,073,672 | 39,000 |
| **RR=0.85** | | | | | | | | | | | | | |
| MRI-first | 149 | 57 | 13 | 8 | 30 | 3,693 | 172 | 3,865 | 507 | 16,976 | 14,212 | 1,566,021 | >200,000 |
| *Polygenic risk-stratified screening* | | | | | | | | | | | | | |
| 10th centile | 148 | 56 | 12 | 7 | 30 | 3,313 | 172 | 3,485 | 467 | 16,976 | 14,213 | 1,549,087 | >200,000 |
| 20th centile | 146 | 54 | 12 | 7 | 30 | 2,934 | 173 | 3,108 | 427 | 16,976 | 14,213 | 1,473,251 | >200,000 |
| 30th centile | 145 | 51 | 11 | 7 | 30 | 2,557 | 176 | 2,733 | 389 | 16,976 | 14,214 | 1,398,023 | 162,000 |
| 40th centile | 144 | 48 | 11 | 6 | 30 | 2,182 | 180 | 2,362 | 352 | 16,976 | 14,214 | 1,323,385 | 126,000 |
| 50th centile | 143 | 44 | 10 | 6 | 30 | 1,808 | 184 | 1,993 | 315 | 16,975 | 14,214 | 1,249,437 | 97,000 |
| 60th centile | 141 | 39 | 9 | 5 | 30 | 1,437 | 191 | 1,628 | 280 | 16,975 | 14,214 | 1,176,407 | 74,000 |
| 70th centile | 140 | 34 | 7 | 4 | 31 | 1,068 | 199 | 1,267 | 247 | 16,974 | 14,215 | 1,104,766 | 55,000 |
| 80th centile | 138 | 26 | 6 | 3 | 31 | 702 | 211 | 913 | 216 | 16,972 | 14,215 | 1,035,580 | 39,000 |
| 90th centile | 137 | 16 | 4 | 2 | 32 | 343 | 227 | 570 | 189 | 16,970 | 14,214 | 971,988 | 27,000 |
| PSA, 1ng/ml | 150 | 58 | 14 | 9 | 30 | 1,290 | 173 | 1,463 | 392 | 16,976 | 14,211 | 1,282,377 | >200,000 |
| PSA, 3ng/ml | 150 | 52 | 16 | 8 | 30 | 621 | 184 | 805 | 235 | 16,976 | 14,212 | 1,082,730 | 132,000 |

*Continued on the next page*

**Appendix Table B.4**: Scenario analysis: The impact of varying the relative impact of screening every 4 years from age 55–69 on prostate cancer-specific mortality, continued

| Strategy | Prostate cancers | Screen-detected cancers | Interval cancers | Over-diagnosed cancers | Deaths from prostate cancer | Screening MRI | Diagnostic MRI | Total MRI | Biopsies | Life-years | QALYs | Costs (£) | WTP threshold* |
|---|---|---|---|---|---|---|---|---|---|---|---|---|---|
| **RR=0.9** | | | | | | | | | | | | | |
| MRI-first | 149 | 57 | 13 | 8 | 32 | 3,693 | 172 | 3,865 | 507 | 16,971 | 14,209 | 1,575,075 | >200,000 |
| *Polygenic risk-stratified screening* | | | | | | | | | | | | | |
| 10th centile | 148 | 56 | 12 | 7 | 32 | 3,313 | 172 | 3,485 | 467 | 16,971 | 14,209 | 1,557,948 | >200,000 |
| 20th centile | 146 | 54 | 12 | 7 | 32 | 2,934 | 173 | 3,108 | 427 | 16,972 | 14,210 | 1,481,808 | >200,000 |
| 30th centile | 145 | 51 | 11 | 7 | 32 | 2,557 | 176 | 2,733 | 389 | 16,972 | 14,211 | 1,406,175 | >200,000 |
| 40th centile | 144 | 48 | 11 | 6 | 32 | 2,182 | 180 | 2,362 | 352 | 16,972 | 14,211 | 1,331,031 | >200,000 |
| 50th centile | 143 | 44 | 10 | 6 | 32 | 1,808 | 184 | 1,993 | 315 | 16,972 | 14,212 | 1,256,459 | >200,000 |
| 60th centile | 141 | 39 | 9 | 5 | 32 | 1,437 | 191 | 1,628 | 280 | 16,971 | 14,212 | 1,182,668 | 181,000 |
| 70th centile | 140 | 34 | 7 | 4 | 32 | 1,068 | 199 | 1,267 | 247 | 16,971 | 14,213 | 1,110,092 | 108,000 |
| 80th centile | 138 | 26 | 6 | 3 | 32 | 702 | 211 | 913 | 216 | 16,970 | 14,213 | 1,039,730 | 65,000 |
| 90th centile | 137 | 16 | 4 | 2 | 32 | 343 | 227 | 570 | 189 | 16,969 | 14,213 | 974,576 | 38,000 |
| PSA, 1ng/ml | 150 | 58 | 14 | 9 | 32 | 1,290 | 173 | 1,463 | 392 | 16,971 | 14,208 | 1,291,400 | >200,000 |
| PSA, 3ng/ml | 150 | 52 | 16 | 8 | 32 | 621 | 184 | 805 | 235 | 16,971 | 14,209 | 1,091,776 | >200,000 |

Scenario analyses in which the age at which screening starts and stops is varied. All analyses follow 1,000 individuals from age 50 to 89 with screening every 4 years, but with different starting and stopping ages. Note that in baseline analyses, screening is from 55-69; this is repeated here but having run the model from age 50. MRI-first refers to MRI-first screening and PSA refers to risk stratification with PSA prior to an MRI.

* Willingness-to-pay (WTP) threshold needed for the strategy to have an incremental net health benefit (NHB) greater than that of no screening. For context, the UK National Institute for Health and Care Excellence set thresholds for most interventions at between £25,000–£35,000 to be considered cost-effective.

Abbreviations: MRI, magnetic resonance imaging; QALYs, quality-adjusted life-years; RR, relative reduction

**Appendix Table B.5:** Scenario analysis: The impact of varying the mean sojourn time (MST) of a screening programme from age 55–69 every 4 years

| Strategy | Prostate cancers | Screen-detected cancers | Interval cancers | Over-diagnosed cancers | Deaths from prostate cancer | Screening MRI | Diagnostic MRI | Total MRI | Biopsies | Life-years | QALYs | Costs (£) | WTP threshold* |
|---|---|---|---|---|---|---|---|---|---|---|---|---|---|
| No screening | 140 | 0 | 0 | 0 | 34 | 0 | 263 | 263 | 176 | 16,965 | 14,211 | 878,516 | N/A |
| **MST=7** | | | | | | | | | | | | | |
| MRI-first | 150 | 55 | 13 | 6 | 28 | 3,694 | 179 | 3,872 | 511 | 16,982 | 14,217 | 1,564,325 | 105,000 |
| *Polygenic risk-stratified screening* | | | | | | | | | | | | | |
| 10th centile | 149 | 54 | 13 | 6 | 28 | 3,314 | 179 | 3,492 | 471 | 16,982 | 14,217 | 1,547,282 | 97,000 |
| 20th centile | 148 | 52 | 12 | 6 | 28 | 2,935 | 180 | 3,115 | 432 | 16,982 | 14,218 | 1,471,359 | 83,000 |
| 30th centile | 147 | 50 | 12 | 6 | 28 | 2,558 | 182 | 2,740 | 393 | 16,982 | 14,218 | 1,396,061 | 70,000 |
| 40th centile | 145 | 47 | 11 | 5 | 28 | 2,183 | 185 | 2,368 | 355 | 16,981 | 14,218 | 1,321,376 | 59,000 |
| 50th centile | 144 | 43 | 10 | 5 | 29 | 1,809 | 189 | 1,998 | 319 | 16,980 | 14,218 | 1,247,407 | 50,000 |
| 60th centile | 142 | 38 | 9 | 4 | 29 | 1,437 | 195 | 1,632 | 283 | 16,979 | 14,218 | 1,174,397 | 41,000 |
| 70th centile | 141 | 33 | 8 | 4 | 29 | 1,068 | 203 | 1,271 | 249 | 16,978 | 14,217 | 1,102,837 | 32,000 |
| 80th centile | 139 | 25 | 6 | 3 | 30 | 703 | 213 | 916 | 217 | 16,976 | 14,217 | 1,033,837 | 25,000 |
| 90th centile | 138 | 16 | 4 | 2 | 31 | 343 | 229 | 572 | 190 | 16,972 | 14,215 | 970,654 | 19,000 |
| PSA, 1ng/ml | 150 | 52 | 15 | 6 | 29 | 1,294 | 184 | 1,478 | 400 | 16,981 | 14,217 | 1,270,278 | 64,000 |
| PSA, 3ng/ml | 149 | 45 | 17 | 5 | 29 | 623 | 194 | 817 | 242 | 16,980 | 14,216 | 1,070,349 | 33,000 |
| **MST=8** | | | | | | | | | | | | | |
| MRI-first | 153 | 61 | 12 | 8 | 28 | 3,682 | 172 | 3,854 | 506 | 16,981 | 14,214 | 1,579,019 | 185,000 |
| *Polygenic risk-stratified screening* | | | | | | | | | | | | | |
| 10th centile | 152 | 60 | 12 | 8 | 28 | 3,302 | 172 | 3,474 | 465 | 16,982 | 14,215 | 1,561,554 | 161,000 |
| 20th centile | 151 | 58 | 11 | 8 | 28 | 2,924 | 174 | 3,097 | 426 | 16,981 | 14,215 | 1,485,026 | 131,000 |
| 30th centile | 149 | 55 | 11 | 7 | 28 | 2,547 | 176 | 2,723 | 388 | 16,981 | 14,215 | 1,408,963 | 107,000 |
| 40th centile | 148 | 52 | 10 | 7 | 28 | 2,173 | 180 | 2,352 | 351 | 16,980 | 14,216 | 1,333,346 | 87,000 |
| 50th centile | 146 | 48 | 9 | 6 | 28 | 1,800 | 184 | 1,984 | 314 | 16,980 | 14,216 | 1,258,262 | 70,000 |
| 60th centile | 144 | 42 | 8 | 6 | 29 | 1,429 | 191 | 1,620 | 279 | 16,979 | 14,216 | 1,183,921 | 55,000 |
| 70th centile | 142 | 36 | 7 | 5 | 29 | 1,061 | 199 | 1,260 | 246 | 16,977 | 14,216 | 1,110,770 | 43,000 |
| 80th centile | 140 | 28 | 5 | 4 | 30 | 697 | 210 | 907 | 215 | 16,975 | 14,216 | 1,039,834 | 31,000 |
| 90th centile | 138 | 18 | 3 | 2 | 31 | 340 | 227 | 567 | 188 | 16,972 | 14,215 | 974,199 | 23,000 |
| PSA, 1ng/ml | 153 | 58 | 14 | 8 | 29 | 1,290 | 179 | 1,468 | 396 | 16,980 | 14,214 | 1,286,062 | 118,000 |
| PSA, 3ng/ml | 152 | 51 | 17 | 7 | 29 | 621 | 189 | 810 | 238 | 16,979 | 14,214 | 1,086,430 | 65,000 |
| **MST=9** | | | | | | | | | | | | | |
| MRI-first | 156 | 67 | 11 | 10 | 28 | 3,669 | 166 | 3,835 | 500 | 16,980 | 14,211 | 1,596,049 | >200,000 |

*Continued on the next page*

**Appendix Table B.5**: Scenario analysis: The impact of varying the mean sojourn time (MST) of a screening programme from age 55–69 every 4 years, continued

| Strategy | Prostate cancers | Screen-detected cancers | Interval cancers | Over-diagnosed cancers | Deaths from prostate cancer | Screening MRI | Diagnostic MRI | Total MRI | Biopsies | Life-years | QALYs | Costs (£) | WTP threshold* |
|---|---|---|---|---|---|---|---|---|---|---|---|---|---|
| *Polygenic risk-stratified screening* | | | | | | | | | | | | | |
| 10th centile | 155 | 66 | 11 | 10 | 28 | 3,289 | 166 | 3,456 | 460 | 16,980 | 14,212 | 1,578,105 | >200,000 |
| 20th centile | 153 | 64 | 11 | 10 | 28 | 2,912 | 168 | 3,080 | 421 | 16,980 | 14,212 | 1,500,887 | >200,000 |
| 30th centile | 152 | 61 | 10 | 9 | 28 | 2,536 | 170 | 2,706 | 383 | 16,980 | 14,213 | 1,423,947 | >200,000 |
| 40th centile | 150 | 57 | 10 | 9 | 28 | 2,162 | 174 | 2,336 | 346 | 16,979 | 14,213 | 1,347,261 | 177,000 |
| 50th centile | 148 | 52 | 9 | 8 | 28 | 1,790 | 179 | 1,969 | 310 | 16,979 | 14,214 | 1,270,893 | 128,000 |
| 60th centile | 146 | 47 | 8 | 7 | 29 | 1,420 | 186 | 1,606 | 275 | 16,978 | 14,214 | 1,195,020 | 92,000 |
| 70th centile | 144 | 40 | 7 | 6 | 29 | 1,054 | 195 | 1,249 | 242 | 16,977 | 14,214 | 1,120,031 | 65,000 |
| 80th centile | 141 | 31 | 5 | 5 | 30 | 691 | 207 | 899 | 212 | 16,975 | 14,214 | 1,046,852 | 44,000 |
| 90th centile | 139 | 19 | 3 | 3 | 31 | 336 | 225 | 561 | 186 | 16,972 | 14,214 | 978,369 | 29,000 |
| PSA, 1ng/ml | 156 | 63 | 13 | 10 | 29 | 1,286 | 173 | 1,459 | 391 | 16,979 | 14,211 | 1,303,907 | >200,000 |
| PSA, 3ng/ml | 154 | 56 | 16 | 9 | 29 | 619 | 184 | 803 | 235 | 16,977 | 14,211 | 1,104,099 | >200,000 |
| **MST=10** | | | | | | | | | | | | | |
| MRI-first | 159 | 74 | 11 | 13 | 28 | 3,655 | 160 | 3,816 | 495 | 16,979 | 14,208 | 1,615,268 | >200,000 |
| *Polygenic risk-stratified screening* | | | | | | | | | | | | | |
| 10th centile | 158 | 72 | 11 | 13 | 28 | 3,276 | 161 | 3,437 | 455 | 16,979 | 14,209 | 1,596,792 | >200,000 |
| 20th centile | 156 | 70 | 10 | 12 | 28 | 2,899 | 162 | 3,061 | 416 | 16,979 | 14,209 | 1,518,802 | >200,000 |
| 30th centile | 154 | 66 | 10 | 12 | 28 | 2,524 | 165 | 2,689 | 378 | 16,979 | 14,210 | 1,440,881 | >200,000 |
| 40th centile | 152 | 62 | 9 | 11 | 28 | 2,150 | 169 | 2,320 | 341 | 16,978 | 14,210 | 1,362,994 | >200,000 |
| 50th centile | 150 | 57 | 8 | 10 | 28 | 1,779 | 175 | 1,954 | 306 | 16,978 | 14,211 | 1,285,185 | >200,000 |
| 60th centile | 148 | 51 | 7 | 9 | 29 | 1,411 | 182 | 1,593 | 272 | 16,977 | 14,212 | 1,207,590 | >200,000 |
| 70th centile | 145 | 43 | 6 | 7 | 29 | 1,046 | 192 | 1,237 | 239 | 16,976 | 14,212 | 1,130,530 | 143,000 |
| 80th centile | 143 | 34 | 5 | 6 | 30 | 685 | 204 | 890 | 210 | 16,974 | 14,213 | 1,054,822 | 76,000 |
| 90th centile | 140 | 21 | 3 | 4 | 31 | 332 | 223 | 556 | 185 | 16,972 | 14,213 | 983,118 | 41,000 |
| PSA, 1ng/ml | 159 | 69 | 13 | 12 | 29 | 1,281 | 168 | 1,449 | 386 | 16,978 | 14,208 | 1,323,679 | >200,000 |
| PSA, 3ng/ml | 157 | 62 | 16 | 11 | 29 | 617 | 180 | 797 | 232 | 16,976 | 14,208 | 1,123,234 | >200,000 |

Scenario analyses in which the mean sojourn time was set to values between 7 and 10. All strategies followed 1,000 individuals from age 55 through 89. Where relevant, screening was every 4 years between 55 and 69.

* Willingness-to-pay (WTP) threshold needed for the strategy to have an incremental net health benefit (NHB) greater than that of no screening. For context, the UK National Institute for Health and Care Excellence set thresholds for most interventions at between £25,000–£35,000 to be considered cost-effective. MRI-first refers to MRI-first screening and PSA refers to PSA followed by reflex MRI.

Abbreviations: MRI, magnetic resonance imaging; QALYs, quality-adjusted life-years; PSA, prostate specific antigen.

**Appendix Table B.6:** Scenario analysis: The impact of varying the age at which screening starts and stops amongst cohorts of 1,000 men followed from age 50–89 years

| Strategy | Prostate cancers | Screen-detected cancers | Interval cancers | Over-diagnosed cancers | Deaths from prostate cancer | Screening MRI | Diagnostic MRI | Total MRI | Biopsies | Life-years | QALYs | Costs (£) | WTP threshold* |
|---|---|---|---|---|---|---|---|---|---|---|---|---|---|
| No screening | 142 | 0 | 0 | 0 | 33 | 0 | 266 | 266 | 178 | 18,665 | 15,735 | 760,164 | N/A |
| **Screening: 50-69** | | | | | | | | | | | | | |
| MRI-first | 150 | 54 | 12 | 7 | 27 | 4,658 | 179 | 4,837 | 614 | 18,682 | 15,743 | 1,572,307 | 109,000 |
| *Polygenic risk-stratified screening* | | | | | | | | | | | | | |
| 10th centile | 149 | 53 | 12 | 7 | 27 | 4,182 | 179 | 4,361 | 563 | 18,682 | 15,743 | 1,543,608 | 102,000 |
| 20th centile | 147 | 51 | 12 | 6 | 27 | 3,707 | 180 | 3,888 | 514 | 18,682 | 15,743 | 1,455,759 | 89,000 |
| 30th centile | 146 | 49 | 11 | 6 | 27 | 3,234 | 183 | 3,417 | 465 | 18,681 | 15,743 | 1,368,530 | 77,000 |
| 40th centile | 145 | 46 | 10 | 6 | 27 | 2,762 | 186 | 2,948 | 417 | 18,681 | 15,743 | 1,281,914 | 66,000 |
| 50th centile | 144 | 42 | 9 | 5 | 28 | 2,292 | 190 | 2,483 | 371 | 18,680 | 15,743 | 1,196,008 | 57,000 |
| 60th centile | 142 | 38 | 8 | 5 | 28 | 1,824 | 196 | 2,021 | 325 | 18,679 | 15,743 | 1,111,032 | 47,000 |
| 70th centile | 141 | 32 | 7 | 4 | 28 | 1,359 | 204 | 1,563 | 281 | 18,677 | 15,742 | 1,027,423 | 39,000 |
| 80th centile | 140 | 25 | 6 | 3 | 29 | 897 | 215 | 1,112 | 239 | 18,675 | 15,741 | 946,154 | 31,000 |
| 90th centile | 139 | 16 | 3 | 2 | 30 | 440 | 231 | 671 | 201 | 18,672 | 15,740 | 870,030 | 24,000 |
| PSA, 1ng/ml | 151 | 55 | 13 | 8 | 27 | 1,628 | 180 | 1,808 | 469 | 18,680 | 15,741 | 1,229,692 | 83,000 |
| PSA, 3ng/ml | 151 | 49 | 16 | 7 | 28 | 783 | 191 | 974 | 269 | 18,678 | 15,740 | 998,589 | 54,000 |
| **Screening: 50-74** | | | | | | | | | | | | | |
| MRI-first | 153 | 88 | 25 | 16 | 26 | 6,117 | 123 | 6,239 | 731 | 18,683 | 15,741 | 1,716,011 | 155,000 |
| *Polygenic risk-stratified screening* | | | | | | | | | | | | | |
| 10th centile | 152 | 86 | 25 | 16 | 26 | 5,482 | 124 | 5,606 | 664 | 18,683 | 15,741 | 1,670,580 | 140,000 |
| 20th centile | 151 | 83 | 24 | 15 | 26 | 4,850 | 128 | 4,978 | 600 | 18,683 | 15,742 | 1,566,397 | 121,000 |
| 30th centile | 149 | 79 | 23 | 15 | 26 | 4,222 | 133 | 4,355 | 537 | 18,682 | 15,742 | 1,463,181 | 104,000 |
| 40th centile | 148 | 74 | 21 | 14 | 27 | 3,598 | 140 | 3,737 | 475 | 18,682 | 15,742 | 1,360,942 | 88,000 |
| 50th centile | 146 | 67 | 19 | 12 | 27 | 2,977 | 148 | 3,125 | 415 | 18,681 | 15,742 | 1,259,816 | 74,000 |
| 60th centile | 145 | 60 | 17 | 11 | 27 | 2,360 | 159 | 2,520 | 357 | 18,679 | 15,742 | 1,160,093 | 61,000 |
| 70th centile | 143 | 51 | 14 | 9 | 28 | 1,750 | 173 | 1,923 | 302 | 18,678 | 15,741 | 1,062,324 | 49,000 |
| 80th centile | 141 | 39 | 11 | 7 | 29 | 1,147 | 191 | 1,338 | 250 | 18,675 | 15,741 | 967,681 | 37,000 |
| 90th centile | 140 | 24 | 7 | 4 | 30 | 557 | 217 | 774 | 204 | 18,672 | 15,739 | 879,389 | 28,000 |
| PSA, 1ng/ml | 156 | 88 | 28 | 18 | 27 | 2,137 | 126 | 2,264 | 542 | 18,681 | 15,738 | 1,314,420 | 163,000 |
| PSA, 3ng/ml | 156 | 80 | 33 | 17 | 28 | 1,028 | 143 | 1,171 | 281 | 18,679 | 15,737 | 1,040,333 | 119,000 |

*Continued on the next page*

**Appendix Table B.6**: Scenario analysis: The impact of varying the age at which MRI-first screening starts and stops amongst cohorts of 1,000 men followed from age 50–89 years, continued

| Strategy | Prostate cancers | Screen-detected cancers | Interval cancers | Over-diagnosed cancers | Deaths from prostate cancer | Screening MRI | Diagnostic MRI | Total MRI | Biopsies | Life-years | QALYs | Costs (£) | WTP threshold* |
|---|---|---|---|---|---|---|---|---|---|---|---|---|---|
| **Screening: 55-69** | | | | | | | | | | | | | |
| MRI-first | 150 | 56 | 12 | 7 | 27 | 3,618 | 176 | 3,794 | 502 | 18,681 | 15,742 | 1,314,427 | 81,000 |
| *Polygenic risk-stratified screening* | | | | | | | | | | | | | |
| 10th centile | 149 | 55 | 12 | 7 | 27 | 3,245 | 176 | 3,422 | 462 | 18,681 | 15,742 | 1,311,425 | 78,000 |
| 20th centile | 148 | 53 | 12 | 7 | 27 | 2,874 | 178 | 3,052 | 424 | 18,680 | 15,742 | 1,249,297 | 67,000 |
| 30th centile | 146 | 50 | 11 | 7 | 27 | 2,505 | 180 | 2,685 | 386 | 18,680 | 15,742 | 1,187,814 | 59,000 |
| 40th centile | 145 | 47 | 10 | 6 | 27 | 2,137 | 184 | 2,321 | 350 | 18,679 | 15,742 | 1,126,970 | 50,000 |
| 50th centile | 144 | 43 | 9 | 6 | 28 | 1,771 | 188 | 1,959 | 314 | 18,679 | 15,742 | 1,066,865 | 43,000 |
| 60th centile | 143 | 39 | 8 | 5 | 28 | 1,407 | 195 | 1,601 | 280 | 18,678 | 15,742 | 1,007,719 | 36,000 |
| 70th centile | 141 | 33 | 7 | 4 | 29 | 1,045 | 203 | 1,248 | 247 | 18,676 | 15,742 | 949,973 | 30,000 |
| 80th centile | 140 | 26 | 6 | 3 | 29 | 687 | 214 | 901 | 216 | 18,674 | 15,741 | 894,599 | 24,000 |
| 90th centile | 139 | 16 | 3 | 2 | 30 | 335 | 230 | 566 | 190 | 18,671 | 15,740 | 844,384 | 20,000 |
| PSA, 1ng/ml | 152 | 57 | 13 | 9 | 27 | 1,264 | 177 | 1,441 | 389 | 18,679 | 15,740 | 1,082,502 | 69,000 |
| PSA, 3ng/ml | 151 | 51 | 16 | 8 | 28 | 609 | 188 | 797 | 236 | 18,677 | 15,739 | 921,491 | 40,000 |
| **Screening: 55-74** | | | | | | | | | | | | | |
| MRI-first | 152 | 73 | 19 | 12 | 27 | 4,371 | 147 | 4,518 | 562 | 18,682 | 15,741 | 1,389,733 | 98,000 |
| *Polygenic risk-stratified screening* | | | | | | | | | | | | | |
| 10th centile | 151 | 72 | 18 | 12 | 27 | 3,917 | 148 | 4,065 | 514 | 18,682 | 15,742 | 1,377,918 | 92,000 |
| 20th centile | 149 | 69 | 18 | 11 | 27 | 3,466 | 150 | 3,616 | 468 | 18,681 | 15,742 | 1,307,192 | 80,000 |
| 30th centile | 148 | 66 | 17 | 11 | 27 | 3,017 | 154 | 3,171 | 423 | 18,681 | 15,742 | 1,237,299 | 69,000 |
| 40th centile | 147 | 62 | 16 | 10 | 27 | 2,570 | 160 | 2,729 | 379 | 18,680 | 15,742 | 1,168,241 | 59,000 |
| 50th centile | 145 | 57 | 14 | 9 | 27 | 2,126 | 167 | 2,292 | 337 | 18,679 | 15,742 | 1,100,140 | 50,000 |
| 60th centile | 144 | 50 | 13 | 8 | 28 | 1,685 | 175 | 1,861 | 296 | 18,678 | 15,742 | 1,033,254 | 41,000 |
| 70th centile | 142 | 43 | 11 | 7 | 28 | 1,249 | 187 | 1,436 | 258 | 18,677 | 15,741 | 968,087 | 34,000 |
| 80th centile | 141 | 33 | 8 | 5 | 29 | 818 | 202 | 1,020 | 222 | 18,675 | 15,741 | 905,718 | 27,000 |
| 90th centile | 139 | 20 | 5 | 3 | 30 | 397 | 223 | 620 | 191 | 18,672 | 15,739 | 849,168 | 21,000 |
| PSA, 1ng/ml | 154 | 74 | 20 | 14 | 27 | 1,527 | 149 | 1,676 | 427 | 18,680 | 15,739 | 1,126,500 | 99,000 |

| | | | | | | | | | | | | | |
|---|---|---|---|---|---|---|---|---|---|---|---|---|---|
| PSA, 3ng/ml | 154 | 67 | 25 | 13 | 28 | 735 | 163 | 899 | 242 | 18,678 | 15,738 | 942,869 | 58,000 |

Scenario analyses in which the age at which screening starts and stops is varied. All analyses follow 1,000 individuals from age 50 to 89 with MRI-first screening every 4 years, but with different starting and stopping ages. Note that in baseline analyses, screening is from 55-69; this is repeated here but having run the model from age 50. MRI-first refers to MRI-first screening and PSA refers to PSA prior to a reflex MRI.

* Willingness-to-pay (WTP) threshold needed for the strategy to have an incremental net health benefit (NHB) greater than that of no screening. For context, the UK National Institute for Health and Care Excellence set thresholds for most interventions at between £25,000–£35,000 to be considered cost-effective.

Abbreviations: MRI, magnetic resonance imaging; QALYs, quality-adjusted life-years; RR, relative reduction; PSA, prostate specific antigen.

**Appendix Table B.7:** Outcomes per 1,000 men for age-based and risk-based MRI-first screening (PI-RADS $\geq$ 3)

| Strategy | Prostate cancers | Screen-detected cancers | Interval cancers | Over-diagnosed cancers | Deaths from prostate cancer | bpMRI | mpMRI | Total MRI | Biopsies | Life-years | QALYs | Costs (£) | WTP threshold* |
|---|---|---|---|---|---|---|---|---|---|---|---|---|---|
| No screening | 140 | 0 | 0 | 0 | 34 | 0 | 263 | 263 | 176 | 16,965 | 14,211 | 878,516 | N/A |
| MRI-first screening | 150 | 61 | 12 | 9 | 27 | 3,686 | 167 | 3,853 | 760 | 16,984 | 14,217 | 1,760,271 | 129,000 |
| *Polygenic risk centiles* | | | | | | | | | | | | | |
| 10 | 149 | 59 | 12 | 9 | 27 | 3,306 | 168 | 3,474 | 694 | 16,984 | 14,218 | 1,722,757 | 117,000 |
| 20 | 148 | 57 | 11 | 8 | 27 | 2,928 | 169 | 3,097 | 628 | 16,983 | 14,218 | 1,626,517 | 100,000 |
| 30 | 146 | 55 | 11 | 8 | 27 | 2,551 | 172 | 2,723 | 564 | 16,983 | 14,218 | 1,531,036 | 86,000 |
| 40 | 145 | 51 | 10 | 7 | 28 | 2,176 | 176 | 2,352 | 500 | 16,982 | 14,218 | 1,436,308 | 73,000 |
| 50 | 143 | 47 | 9 | 7 | 28 | 1,803 | 181 | 1,984 | 438 | 16,981 | 14,218 | 1,342,451 | 61,000 |
| 60 | 142 | 42 | 8 | 6 | 28 | 1,432 | 188 | 1,619 | 378 | 16,980 | 14,218 | 1,249,730 | 50,000 |
| 70 | 140 | 36 | 7 | 5 | 29 | 1,064 | 196 | 1,260 | 319 | 16,978 | 14,217 | 1,158,679 | 40,000 |
| 80 | 139 | 28 | 5 | 4 | 30 | 699 | 208 | 908 | 263 | 16,976 | 14,217 | 1,070,475 | 30,000 |
| 90 | 138 | 17 | 3 | 2 | 31 | 341 | 226 | 567 | 211 | 16,973 | 14,215 | 988,512 | 22,000 |
